# RyR1 protein content is reduced in inflammatory and mitochondrial myopathies and induces ER stress

**DOI:** 10.1101/2023.02.01.23285343

**Authors:** Jeremy Vidal, Eric Fernandez, Martin Wohlwend, Pirkka-Pekka Laurila, Andrea Lopez Mejia, Julien Ochala, Isabel Christina Lopez Mejia, Johannes Alexandre Lobrinus, Bengt Kayser, Nicolas Place, Nadège Zanou

## Abstract

Decreased ryanodine receptor type 1 (RyR1) protein is a hallmark of recessive *RYR1*-related myopathies (*RyR1*-RM), which are caused by recessive mutations in the *RYR1* gene. It is not clear how the decrease in the RyR1 protein triggers muscular disorders. Furthermore, it is a hot topic whether a decrease in RyR1 protein levels can also occur during non-*RYR1*-related myopathies. In this study, we first show that reduced *RYR1* transcripts are associated with various human myopathies, and that RyR1 protein levels are significantly decreased in muscle samples analysed in inflammatory myopathies (IM) and mitochondrial myopathies (MM), both of which are non-*RYR1*-RM. Secondly, proteomic data show that exclusive depletion of RyR1 protein *in vitro* recapitulates the common altered molecular pathways observed during myopathies. RyR1 protein depletion impairs ER-mitochondria tethering and Ca^2+^ transfer to mitochondria, decreases mitophagy genes and induces an accumulation of dysfunctional mitochondria. This phenomenon is also associated with altered lipid homeostasis with an increase in deleterious sphingolipid species. Finally, decreased RyR1 protein levels lead to an increase in the ER stress markers GRP78-Bip and CHOP in muscle cell *in vitro*, and in mouse and human muscles. Overall, our results indicate an important role of RyR1 protein depletion and ER stress in the pathogenesis of myopathies.

## Introduction

Myopathies present a variety of symptoms and cases, rending their diagnosis difficult. Any scientific breakthrough establishing credible and early markers of these diseases would be beneficial for the management of patients. RyR1 is the largest known homotetramer involved in the muscle contraction, with a total molecular mass >2 MDa ^1^. This large protein is known to interact with a broad range of other cellular proteins and undergoes numerous post-translational modification (PTM). For instance, the RyR1 channel is stabilized by a protein with a key role in calcium homeostasis—the FKBP12-encoded calstabin1 (for calcium channel stabilizing binding protein 1). PTMs at specific RyR1 residues, such as oxidation, phosphorylation, nitrosylation, and acetylation, lead to calstabin1 dissociation and trigger Ca^2+^ leak from the RyR1. While exercise-induced acute RyR1 Ca^2+^ leak promotes mitochondrial adaptations ^2^, a sustained leaky RyR1 channel has been linked to several pathological conditions, such as myopathy, cancer-induced cachexia, or ageing-induced sarcopenia ^3-5^.

Apart from PTMs and calstabin1 binding, the function of RyR1 can also be altered through mutations, which trigger the so-called the RyR1-related myopathies (*RYR1*-RMs). These mutations can be either homozygous or heterozygous and lead to distinct channel defects ranging from leaky RyR1, E-C uncoupling (not responsive to electrical or agonist stimulations), to loss of RyR1 protein, the latter being caused by recessive RyR1 mutations. *RYR1-*RMs constitute the most common class of congenital myopathies, including central core disease, multicore disease, core-rod myopathy, centronuclear myopathy, and congenital fiber-type disproportion ^6 7 8^. Attempts to study how recessive mutations in the *RyR1* gene leading to a decrease in RyR1 protein levels induce myopathies have proved difficult. Indeed, complete deletion of RyR1 *in vivo* is lethal, making it difficult to study the role of RyR1 protein depletion in the pathogenesis of myopathies.

An inducible muscle specific *RYR1* knock-out mouse model (*RYR1*-Rec) with depletion of 50% of the RyR1 protein, which recapitulates most features of the *RYR1*-RM Dusty core disease has been recently developed ^9^. This model demonstrated that 50% RyR1 protein reduction is sufficient to induce the disease. How the decrease in RyR1 protein triggers these disorders remains however poorly understood and requires further investigations.

The role of RyR1 in myopathies is extensively investigated in *RYR1-RM*, but RyR1 alterations (expression or function or both) might also be associated to other myopathies. Indeed, a previous study has observed a decrease in RyR1 protein levels in inclusion body myositis (IBM), a severe form of inflammatory myopathy (IM) with no mutations on the *RYR1* gene, which has been linked to calpain-induced protein degradation ^10^.

In the present study, combining bioinformatics, proteomics, molecular biology, and imaging, we aimed at investigating whether RyR1 protein decrease might also be associated to non-*RYR1*-related myopathies and how reduced RyR1 content triggers muscular disorders.

Our results showed decreased *RYR1* transcripts in several non-*RYR1*-RM including juvenile dermatomyositis, polymyositis, necrotizing myopathy, IBM, myotonic dystrophy 2 (DM2), Becker and Duchenne Muscular Dystrophy (DMD) at both preclinical and clinical stages and in limb girdle muscular dystrophy type 2 A (LGMD2). Interestingly, RyR1 protein levels were significantly decreased in all tested muscle samples of patients suffering from inflammatory myopathies (IM) and mitochondrial myopathies (MM) as compared to their age-matched controls. RyR1 depletion *in vitro* in myoblasts led to myotubes showing a wide range of disorders, which covered the molecular phenotypes observed in human myopathies. Our molecular investigations revealed that RyR1 depletion induces mitochondrial aggregation and dysfunction through defects in ER-mitochondria tethering, defects in mitophagy and altered lipid homeostasis. These disorders were associated with increased ER stress markers, which we also observed in RyR1-Rec muscles and IM and MM muscle samples. Interestingly, bioinformatic analyses revealed a correlation between the decreased RyR1 transcript levels and increased ER stress markers, suggesting that the decrease in RyR1 content increased ER stress markers, a key feature of myopathies.

## Materials and methods

### Ethical Approval

Our research complies with all relevant ethical regulations; the study was validated by the Commission d’éthique de la recherche sur l’être humain du Canton de Vaud with the reference CER-VD 2019-00356.

### Human muscle biopsies

All the human muscle biopsies were obtained from the Institut Universitaire de Pathologie and the Biobanque of the Canton de Vaud, where a clear diagnosis was established for each patient. All the subjects provided consent in exploiting their sample for research purpose.

### Muscle biopsies examination

Muscle tissue was frozen in isopentane cooled in liquid nitrogen. Cryosections were stained using H&E, special stains and immunohistochemistry.

The main diagnostic criteria for inflammatory myopathies were presence of endo- or perimysial lymphocytic infiltrate, with or without muscle fibers necrosis. For inclusion body myositis (IBM), presence of rimmed vacuoles and TDP43-positive inclusions was required.

The diagnostic criteria for mitochondrial myopathy were presence of ragged-red-fibers and COX-negative muscle fibers.

### Gene expression analysis of muscle diseases

Gene expression data for different muscular dystrophies (control n=18, Becker muscular dystrophy n=5, Duchenne muscular dystrophy n=10, juvenile dermatomyositis n=21, limb girdle muscular dystrophy 2A n=10) were obtained from the publicly available E-GEOD-3307 dataset^11^. Gene expression data for different inflammatory myositis disorders (control n=5, Dermatomyositis n=8, Polymyositis n=8, Necrotizing myositis n=5, Inclusion body myositis n=10) were retrieved from the publicly available GSE39454 dataset ^12^. RYR1 expression levels in skeletal muscle of control (n=10) or DM2 patients (n=10) were obtained from GSE37084 ^13^ whereas RYR1 muscle expression levels of control (n=14) and presymptomatic Duchenne muscular dystrophy children (n=23) were obtained from GSE6011 ^14^. Preprocessed data were used for data analysis. Student’s two-tailed t-tests were used to compare two groups and one-Way ANOVAs followed by Dunnett’s multiple comparisons test were used to compare more than two groups. Heatmaps are shown as log2 fold change of each muscle disease relative to controls. Gene expression correlation from *postmortem* skeletal muscle biopsies were performed using the GTEx project v7, n=491. Pearson correlation and Student p-values were calculated using the corAndPvalue function in WGCNA package, R Bioconductor.

### Mouse model

Inducible RyR1 recessive (RyR1-Rec) mouse was generated as previously reported (Pelletier et al, 2020; Shuler et al, 2015). Briefly, C57BL/6 J background RyR1^flox/flox^ mouse was generated by introducing LoxP sites on both sides of exons 9-11 of the RYR1 gene and was crossed with the HSA-Cre-ER^T2^ mouse line, in which the expression of the tamoxifen dependent Cre-ER^T2^ recombinase is under the control of the human skeletal muscle α-actin gene (Shuler et al 2015). This generates the RyR1^flox/flox^::HSA-Cre-ER^T2^ mouse line. RYR1 gene was then deleted by tamoxifen injection specifically in skeletal muscle fibers. Mice were given *ad libitum* access to food and water, food pellets were provided directly in the cage to ensure a correct feeding.

Two-month male mice were injected with tamoxifen kept for 75 days before sacrifice.

### Cell culture

C2C12 mouse skeletal myoblasts were obtained from the American Type Culture Collection and grown in proliferation medium composed of Dulbecco’s modified Eagle’s medium (DMEM; Thermo Fisher Scientific, Basel, Switzerland) supplemented with 10% fetal bovine serum (Thermo Fisher Scientific, Basel, Switzerland), 100 IU/ml penicillin, 100 μg/ml streptomycin (Thermo Fisher Scientific, Basel, Switzerland) and 1% non-essential amino acids (Thermo Fisher Scientific, Basel, Switzerland), and maintained at 37 °C in a humidified atmosphere with 5% CO_2_. To induce differentiation, myoblasts were grown to 80-90% confluence, the proliferation medium was then replaced with a differentiation medium (DM), consisting of DMEM supplemented with 2% horse serum (Thermo Fisher Scientific, Basel, Switzerland).

### Si-RNA transfection

To perform the si-RNA transfection, ∼150.000 C2C12 myoblasts were plated per well of 6-well plates and incubated for proliferation for 24h. Then the myoblasts were transfected with either 10, 50 or 100 nM final concentration of silencer select negative control si-RNA (si-CTRL) (**4390843**, Thermo Fisher Scientific, Basel, Switzerland) or silencer select pre-designed si-RNA directed against RyR1 (si-RyR1) (B1, **s73237 or B2, s73238** Thermo Fisher Scientific, Basel, Switzerland) using Lipofectamine RNAimax protocol according to the manufacturer (Thermo Fisher Scientific, Basel, Switzerland). The differentiation was induced the day after the transfection and the analyses were performed at day 4 of transfection and 3 of differentiation (D4TD3D) or at day 7 of transfection and day 6 of differentiation (D7TD6D).

### Western blot

A lysis buffer containing the following: 20 mM Tris/HCl (pH 6.8), 2 mM EDTA (pH 8), 137 mM NaCl, 10% glycerol, 1% Triton X-100, 10 mM glycero-phosphate, 1 mM KH_2_PO4, 1 mM PMSF, 1 mM NaVO3, 50 mM NaF, 10 mM NaPPi, and a protease inhibitor mixture (Roche, Complete Mini, Basel, Switzerland) was used to resuspend myotube pellets (100 μl / well of a 6-well plate) or human muscle samples (100 μl / 5 mg of tissue). The preparation was homogenized with pipette tips for cells or potter for muscles, incubated 1 h at 4 °C and gently sonicated. Then, nuclei and debris were removed by centrifugation at 10’000 *g* at 4 °C for 10 min. Protein quantification was assessed using the BCA kit (Thermo Fisher Scientific, Ecublens, Switzerland). Ten to 20 μg of protein were incubated with 2 x Laemmli sample buffer containing SDS and 2-mercapto-ethanol (Bio-Rad, Hercules, CA, USA) for 3 min at 95 °C, electrophoresed 1 h on 4-15% SDS-precast gradient gels (Bio-Rad, Hercules, CA, USA), and wet transferred 1 h onto PVDF membranes. Membranes were stained with Red Ponceau (homemade) and total protein bands were quantified using the Image Studio software v 5.2.5 (LI-COR, Lincoln, NE, USA). Then, the Red Ponceau was washed out with PBS and the membranes were saturated 1h at room temperature with phosphate buffered solution (PBS)-LI-COR blocking buffer (LI-COR, Lincoln, NE, USA). Blots were incubated overnight with rabbit anti-RyR1 (provided by Prof-Marks lab at Columbia University, 1:2500), anti-FKBP12 (Abcam, Cambridge, UK, 1:1000), mouse anti-SERCA1 (Abcam, Cambridge, UK, 1:1000), rabbit anti-SERCA2 (Abcam, Cambridge, UK, 1:1000), mouse anti-total OXPHOS (Abcam, Cambridge, UK, 1:1000), mouse anti tubulin α (Sigma, S^t^ Louis, USA, 1:5000), rabbit anti acetylated tubulin (Sigma, S^t^ Louis, USA, 1:1000), rabbit anti-GAPDH (Sigma, S^t^ Louis, USA, 1:5000), rabbit anti ADFP (perilipine 2, Abcam, Cambridge, UK, 1:1000), rabbit anti-pyruvate dehydrogenase E1 α phospho serine 293 (Abcam, Cambridge, UK, 1:1000), mouse anti-pyruvate dehydrogenase (Abcam, Cambridge, UK), rabbit anti GRP78-Bip (Abcam, Cambridge, UK, 1:1000), rabbit anti CHOP-DDIT3 (Abcam, Cambridge, UK, 1:1000), rabbit anti EIF2 α phospho serine 51 (Abcam, Cambridge, UK, 1:1000), mouse anti EIF2 α (Abcam, Cambridge, UK, 1:1000). Membranes were washed in PBS-buffered saline-Tween 20 (TBS-T) and incubated for 1 h at room temperature with IRDye 680-conjugated donkey anti-mouse or rabbit IgG (LI-COR, Lincoln, NE, USA, 1:10000) and IRDye 800-conjugated donkey anti-mouse or rabbit IgG (LI-COR, Lincoln, NE, USA, 1: 5000) in blocking buffer. Immunoreactive bands were visualized using infrared fluorescence (IR-Odyssey scanner, LI-COR, Lincoln, NE, USA). Band densities were quantified using Image Studio v 5.2.5 (LI-COR, Lincoln, NE, USA). Protein intensity signal was normalized to that total protein staining. The LiCor system allows detection of different proteins at the same time (revealed in different channels). When needed, the membranes were stripped using the appropriate LiCor stripping solution (LiCor, Lincoln, NE, USA). As myopathy samples can undergo several changes, RyR1, SERCA and FKBP12 were normalized to the quantity of myosin (detected using MF20 antibody 1/1000, Life Technologies) to detect their real quantity in muscle cell. Proteins were quantified from at least two different blots, which are then averaged. They were expressed as percentage of si-CTRL for cells and percentage of healthy human muscles for human.

### Real-time polymerase chain reaction

Myotubes were homogenized with 500 μl TRIzol (Invitrogen, Basel, Switzerland) per well of 6-well plates and the extracted RNA was quantified using the Nanodrop (Thermo Fisher Scientific, Basel, Switzerland). One μg RNA was reverse transcribed using qScript Reverse Transcriptase (Invitrogen, Basel, Switzerland). RT-PCR was performed using 2 μl of cDNA, 9 μl of GoTaq qPCR Master Mix (Promega, Dübendorf, Switzerland) and 400 nM of each primer in a total reaction volume of 11 μl. Cyclophilin B housekeeping gene and genes of interest were amplified in parallel. Data were recorded on a ViiA7 real-time RT-PCR detection system (Life Technologies, Basel, Switzerland) and cycle threshold (Ct) values for each reaction were determined using analytical software from the same manufacturer. Each cDNA was amplified in triplicate, and Ct values were averaged for each triplicate. The average Ct value for Cyclophilin B was subtracted from the average Ct value for the gene of interest and normalized to control cells. As amplification efficiencies of the genes of interest and Cyclophilin B were comparable, the amount of mRNA, normalized to Cyclophilin B was given by the relation 2^-ΔΔCt. The sequence of the primers is as follow:

RyR1: Fw, 5’-GAAGGTTCTGGACAAACACGGG-3’ Rev, 5’-TCGCTCTTGTTGTAGAATTTGCGG-3’

MHC Ia: Fw, 5’-AGATCCGAAAGCAACTGGAG-3’ Rev, 5’-CTGCCTTGATCTGGT TGAAC-3’

MHC IIa: Fw, 5’-ATGAGCTCCGACGCCGAG-3’ Rev, 5’-TCTGTTAGCATGAACTGG TAGGCG-3’

Glut-4: Fw, 5’ GATGAGAAACGGAAGTTGGAGAGA Rev, 5’ GCACCACTGCGATGATCAGA

SOD1: Fw, 5’-ATGGGTTCCACGTCCATCAGTA-3’ Rev, 5’-CATTGCCCAGGTCTCCAACA-3’

SOD2; Fw, 5’-GAGAATCTCAGTGCTCACTCGTGTC-3’ Rev, 5’-GGAACCCTAAATGCTGCCAGTC-3’

Catalase: Fw, 5’-CGAGGGTCACGAACTGTGTCA-3’ Rev, 5’-GGTCACCCACGATATCACCAGATAC-3’

BNIP3: Fw, CCTGTCGCAGTTGGGTTC Rev, GAAGTGCAGTTCTACCCAGGAG

PARK2: Fw, CCGAATCACCTGACGGTTCA Rev, TCTGGCTGCTTCTGAATCCC

PINK1: Fw, GCTTGCCAATCCCTTCTATG Rev, CTCTCGCTGGAGCAGTGAC

SQSTM1: Fw, CCTCAGCCCTCTAGGCATTG Rev, TTCTGGGGTAGTGGGTGTCA

BECN1: Fw, CCCAGCCAGGTGCTTACTATT Rev, CATTCCTCACACAGCGGGT

CphB: Fw 5’ TGGAGAGCACCAAGACAGACA Rev, 5’ TGCCGGAGTCGACAATGAT

### Immunofluorescence

Cells prepared in glass bottom MatTek dishes were fixed with 4% paraformaldehyde PBS for 30 min then permeabilized with 0.3% triton diluted in PBS solution for 5 min. The cells were washed with PBS three times for 5 min and blocked with 0.1% triton - 2.5% Donkey serum prepared in PBS solution for 1h30. After aspiration of the blocking buffer, the cells were incubated with the appropriate primary antibody (rabbit acetylated tubulin 1/500, mouse tomm20 1/150) prepared in the blocking buffer for 1h30. The cells were washed three times 5 min with PBS and then incubated 1h30 in the secondary antibodies: Alessa fluor 488-AffiniPure donkey anti rabbit (Jackson ImmunoResearch, 1/500) and Alessa fluor 594-AffiniPure donkey anti mouse (Jackson ImmunoResearch, 1/500) prepared in the blocking buffer and protected from light. After washing the cells three times for 5 min with PBS, the nuclei were stained with Hoechst for 5 min and the Hoechst was washed out three times for 5 min with PBS and replaced by PBS. The acquisitions were performed using a confocal microscope system (Zeiss LSM 5 Live, Oberkochen, Germany; 40x oil immersion lens).

For LDs staining, si-CTRL AND si-RyR1 myotubes at D7TD6D were loaded with the 4,4-Difluoro-1,3,5,7,8-Pentamethyl-4-Bora-3a,4a-Diaza-s-Indacene, BODIPY diluted in the differentiation medium (1/250) and incubated in the incubator for 45 min. The acquisition of the BODIPY signal was performed using the confocal microscope system (Zeiss LSM 5 Live, Oberkochen, Germany; 40x oil immersion lens).

### Cytosolic Ca^2+^ measurements

C2C12 transfected myotubes at D7TD6T were loaded with the cytosolic Ca^2+^ indicator Fluo-4 AM (5μM, Invitrogen, Basel, Switzerland) solubilized in a Krebs solution (in mM: NaCl 135.5, MgCl_2_ 1.2, KCl 5.9, glucose 11.5, HEPES 11.5, CaCl_2_ 1.8, final pH 7.3) for 20 min in the incubator. Cells were then rinsed twice with a Ca^2+^-free Krebs solution (in mM: NaCl 135.5, MgCl_2_ 1.2, KCl 5.9, glucose 11.5, HEPES 11.5, 200 μM Na-EGTA, final pH 7.3). Fluo-4 fluorescence was monitored using a confocal microscope system (Zeiss LSM 5 Live, Oberkochen, Germany; 40x oil immersion lens; the excitation wavelength was 488 nm and the emitted fluorescence was recorded between 495-525 nm). After recording the basal fluorescence, myotubes were stimulated with 2.5 mM (final concentration) caffeine to trigger Ca^2+^ release from the ER. The same cells were then stimulated with 1 μM thapsigargin to inhibit the SERCA pumps and trigger Ca^2+^ leak through the ER. The Zen software (2012 version, Zeiss, Oberkochen, Germany) was used for the acquisition and data were exported to excel files for analyses. The use of the single excitation/emission Fluo-4 dye necessitated normalizing to pre-stimulation values to account for possible differences in dye loading and excitation strength.

### Mitochondrial Ca^2+^ measurements

To investigate mitochondrial Ca^2+^ uptake, si-CTRL and si-RyR1 myotubes at D7TD6D plated in MatTek dishes with a bottom coverslip (MatTek, Ashland, MA, USA) were loaded with 1 ml of Krebs solution containing 1 μM of the mitochondrial fluorescent indicator Rhod-2 AM (Invitrogen, Basel, Switzerland) for 1 h at room temperature. Cells were then washed twice with Krebs solution (mM: NaCl 135.5, MgCl_2_ 1.2, KCl 5.9, glucose 11.5, HEPES 11.5, CaCl_2_ 1.8, final pH 7.3) and Rhod-2 fluorescence was measured using a confocal microscope (inverted Zeiss LSM 710 confocal microscope, Oberkochen, Germany) and 40x oil immersion lens, with excitation at 532 nm and the emitted signal collected through a band pass filter (540-625 nm). To avoid the contribution of RyR1 in mitochondrial Ca^2+^ uptake in si-CTRL myotubes, all the dishes were pre-treated with 10 μM final concentration of dantrolene. After recording the basal fluorescence, myotubes were stimulated with 1 μM final concentration of thapsigargin and change in Rhod-2 fluorescence was calculated by reporting the peak of fluorescence to the baseline (normalized fluorescence).

### Mitochondrial O_2_ flux measurements using high resolution respirometry

D7TD6D transfected myotubes were used to measure mitochondrial respiration as previously reported ^2^. The Oroboros O2k chambers (Oroboros, Innsbruck, Austria) were equilibrated using the MIRO5 buffer (mM: EGTA 0.5, MgCl_2_ 3, lactobionic acid 60, taurine 20, KH_2_PO_4_ 10, HEPES 20, D-sucrose 110, BSA fatty acid free 1 g / l pH 7.3). Then one well of a 6-well plate was trypsinized 3 min with Trypsin / EDTA 0.05% (Invitrogen, Basel, Switzerland), centrifuged for 5 min at room temperature and 1000 rpm, then gently washed with a MIRO5 buffer and resuspended in 2.5 ml of MiRO5 solution and 2 mL of cell suspension inserted in each chamber. Mitochondrial respiration rates were assessed at 37ºC using a slightly modified SUIT-008 protocol of the DatLab 7.3 software (Oroboros, Innsbruck, Austria) (Figure S2p). After stabilization of respiration, the cells were permeabilized using 1 μM digitonin (50 μg / ml), then successively incubated with 5 μl malate (2 mM), 10 μl pyruvate (10 mM) to check leak respiration (MP_L_). MgCl_2_ (0.5 mol/mol ADP) associated to 20 μl ADP (500 mM) was then added to assess OXPHOS capacity supported by pyruvate and malate completed with 5 μl glutamate (10 mM), N-linked pathway, (N_P_). The integrity of the mitochondria was checked after addition of ADP by using 5 μl cytochrome C (4 mM). To investigate the activity of the N- and succinate-linked pathways (NS_P_), 20 μl of succinate (10 mM) was added to the chamber. The electron transfer system (NS_E_) was assessed by using a 1 μl-titration protocol with FCCP (1 mM); then the activity of complex I was inhibited by 1 μl of rotenone (1 mM) (S_E_) and residual oxygen consumption (ROX) was determined by addition of 1 μl of antimycinin A (5 mM). An air calibration was performed on each experimental day. Protein quantification was performed in each condition on a representative well using the BCA assay (Thermo Fisher Scientific, Basel, Switzerland). The specific O_2_ flux per mass was obtained after normalization of ROX-corrected O_2_ fluxes by mg of protein in the chamber.

### Proteomics

Proteomic analysis was performed on cells according to a previously published protocol ^2^.The following protocol applies for human muscle biopsies analysis.

### Tissue extraction

Tissue samples were weighted (8.9-22mg), resuspended in 80% methanol at -20°C (v/v, 150 μl / 10 mg tissue) and homogenized by agitation with ceramic beads twice for 30s in a FastPrep FP120 homogenizer at maximum speed. Samples were cooled on ice for two min between homogenization steps. The solvent was removed, and the homogenized material was allowed to dry under air flow 5 min in the homogenization tubes in a laminar flow hood. The pellet was resuspended in lysis buffer (1% Sodium deoxycholate, 30mM Tris pH 8.6, 10 mM DTT) with a ratio of 150 μl/ 10 mg of initial tissue weight and shaken again in the FastPrep system for 3x20s, with 3 min cooling on ice in between runs. An aliquot of 300 μl was taken, mixed 1:1 (v/v) with lysis buffer, heated at 95°C for 10 min with agitation and used for all subsequent steps. Protein concentrations were determined by the tryptophane fluorescence method ^15^ and adjusted to 2 mg/ml with miST buffer.

### Protein Digestion

Samples (100 μg in 50 μl) were digested following a modified version of the iST method ^16^ (named miST method). After diluting 1:1 with water, reduced disulfides were alkylated by adding 1/4 vol (25 μl) of 160 mM chloroacetamide (final 32 mM) and incubating at 25°C for 45 min in the dark. Samples were adjusted to 3 mM EDTA and digested with 1 μg Trypsin/LysC mix (Promega #V5073) for 1h at 37°C, followed by a second 1h digestion with a second and identical aliquot of proteases. Completeness of digestion was assessed qualitatively by SDS-PAGE migration (data not shown). To remove sodium deoxycholate, two sample volumes of isopropanol containing 1% TFA were added to the digests, and the samples were desalted on a strong cation exchange (SCX) plate (Oasis MCX; Waters Corp., Milford, MA) by centrifugation. After washing with excess isopropanol/1%TFA, peptides were eluted in 250 μl of 80% MeCN, 19% water, 1% (v/v) ammonia, frozen and dried by evaporation.

### Peptide fractionation for library construction

Samples were redissolved in 0.05% trifluroacetic acid, 2% (v/v) acetonitrile for LC-MS/MS analysis. Aliquots of 5 μg of all samples were mixed to create a pool, which was manually separated into 7 fractions by off-line basic reversed-phase (bRP) using the Pierce High pH Reversed-Phase Peptide Fractionation Kit (Thermo Fisher Scientific). The fractions collected were: flow through, 7.5, 10, 12.5, 15, 17.5 and 50% MeCN in 0.1 % triethylamine (∼pH 10). Dried bRP fractions were redissolved in 20 μl buffer A and 2 μl were injected for LC-MS/MS analysis.

### Liquid Chromatography-Mass spectrometry

LC-MS/MS analysis was carried out on a TIMS-TOF Pro (Bruker, Bremen, Germany) mass spectrometer interfaced through a nanospray ion source (“captive spray”) to an Ultimate 3000 RSLCnano HPLC system (Dionex). Peptides were separated on a reversed-phase custom packed 40 cm C18 column (75 μm ID, 100Å, Reprosil Pur 1.9 um particles, Dr. Maisch, Germany) at a flow rate of 0.250 μl/min with a 6-27% acetonitrile gradient in 92 min followed by a ramp to 45% in 15 min and to 95% in 5 min (all solvents contained 0.1% formic acid). Identical LC gradients were used for data dependent-(DDA) and data independent acquisition (DIA) measurements.

For creation of the spectral library, data-dependent acquisition (DDA) was carried out on the 7 bRP fractions using a standard TIMS PASEF method (Meier et al 2018) with ion accumulation for 100 ms for each the survey MS1 scan and the TIMS-coupled MS2 scans. Duty cycle was kept at 100%. Up to 10 precursors were targeted per TIMS scan. Precursor isolation was done with a 2 Th or 3 Th windows below or above m/z 800, respectively. The minimum threshold intensity for precursor selection was 2500. If the inclusion list allowed it, precursors were targeted more than one time to reach a minimum target total intensity of 20’000. Collision energy was ramped linearly based uniquely on the 1/k0 values from 20 (at 1/k0=0.6) to 59 eV (at 1/k0=1.6). Total duration of a scan cycle including one survey and 10 MS2 TIMS scans was 1.16 s. Precursors could be targeted again in subsequent cycles if their signal increased by a factor 4.0 or more. After selection in one cycle, precursors were excluded from further selection for 60s. Mass resolution in all MS measurements was approximately 35’000.

The diaPASEF method used the same instrument parameters as the DDA methods and was as reported previously (Meier et al 2020). Per cycle, the mass range 400-1200 m/z was covered by a total of 32 windows, each 26 Th wide (overlap 1 m/z) and a 1/k0 range of 0.3. Collision energy and resolution settings were the same as in the DDA method. Two windows were acquired per TIMS scan (100ms) so that the total cycle time was 1.7 s.

### Library creation

Raw Bruker MS data were processed directly with Spectronaut 14.10 (Biognosys, Schlieren, Switzerland). A library was constructed from the DDA data for the fractions by searching the reference mouse proteome (RefProt, www.UNIPROT.org) database of November 5^th^, 2019 (55’408 sequences). For identification, peptides of 7-52 AA length were considered, cleaved with Trypsin/P specificity and a maximum of 2 missed cleavages. Carbamidomethyl-Cys (fixed), Met oxidation and N-terminal protein acetylation (variable) were the modifications applied. Mass calibration was dynamic and based on a first database search. The Pulsar engine was used for peptide identification. Protein inference was performed with the IDPicker algorithm. PSM, peptide and protein identifications were all filtered at 1% FDR against a decoy database.

Specific filtering for library construction filtered out fragments corresponding to less than 3 AA and fragments outside the 300-1800 m/z range. Also, only fragments with a minimum base peak intensity of 5% were kept. Precursors with less than 3 fragments were also eliminated and only the best 6 fragments were kept per precursor. For quantitation only PSM’s with maximum one missed cleavage were used. No filtering was done on the basis of charge state. Shared (non proteotypic) peptides were kept.

The library created contained 42’399 precursors mapping to 31’607 peptide sequences, of which 11’193 were proteotypic. These corresponded to 3’648 protein groups (6’243 proteins). Of these, 777 were single hits (one PSM). In total 248’194 fragments were used for quantitation.

### DIA quantitation

Peptide-Centric analysis of DIA data was done with Spectronaut 14.10 using the library described above. Both MS1 and MS2 data were used for quantitation ^17^. Run alignment was assisted by a deep learning algorithm based on sample-specific in silico prediction of retention times of identified peptides and alignment based on local nonlinear regression. Interference correction (from neighboring isotope envelopes) was performed at both the MS1 and MS2 levels using windows of 2 and 3 min respectively. Single hits proteins (defined as matched by one stripped sequence only) were kept in the Spectronaut analysis. Peptide quantitation was based on XIC area, for which a minimum of 1 and a maximum of 3 (the 3 best) precursors were considered for each peptide, from which the median value was selected. Peptides were retained for calculation of protein group quantities if they passed the set Q-value threshold of identification (0.01) in at least 50% of the runs. For the retained peptides, any missing values were imputed with low-shifted values based on the global distribution of values in the entire experiment. Quantities for protein groups were obtained by summing all assigned peptide intensities after filtering. Global normalization of runs/samples was done based on the median of peptides.

In total, 31’373 precursors in the library were quantitated in the samples with the DIA data (cumulative on all runs). These corresponded to 24’752 peptides, resulting in 5’730 inferred proteins in 3’399 protein groups. Mass tolerances were determined dynamically and were typically between 9 and 11 ppm. Average mass errors in individual samples were between 2.7 and 3.5 ppm in MS1 and MS2, respectively. The average of data points per peak was 8.2 in both MS1 and MS2. Total normalized intensities for Protein Groups were exported to .txt files for further analysis.

### Data processing and statistical tests

All subsequent analysis was done with the Perseus software package ^18^. Intensity values were log2-transformed. After removal of contaminants and assignment to groups, a Student’s t-tests was carried out among all conditions, with Benjamini-Hochberg correction for multiple testing (q-value threshold <.05). The difference of means obtained from the test were used for 1D enrichment analysis on associated GO/KEGG annotation as described (Cox and Mann, 2012). The enrichment analysis was also FDR-filtered (Benjamini-Hochberg, q-val<0.02).

### Raw data deposition

All raw MS data together with MaxQuant output tables are available via the Proteomexchange data repository (www.proteomexchange.org) with the accession *PXD039693.

Reviewer account details:

*Username: *

*Password: *wUSZuUOZ

### Transmission electron microscopy

Cells grown on coverslip were fixed in glutaraldehyde solution (EMS, Hatfield, PA, US) 2.5% in Phosphate Buffer (PB 0.1M pH7.4) (Sigma, St Louis, MO, US) during 1 hour at room temperature (RT). Then they directly postfixed by a fresh mixture of osmium tetroxide 1% (EMS, Hatfield, PA, US) with 1.5% of potassium ferrocyanide (Sigma, St Louis, MO, US) in PB buffer during 1 hours at RT. The samples were then washed three times in distilled water and dehydrated in ethanol solution (Sigma, St Louis, MO, US) at graded concentrations (30%-10min; 70%-10min; 100%-3x10min). This was followed by infiltration in Epon (Sigma, St Louis, MO, US) overnight, then replaced the day after with fresh resin in a gelatin capsule which was placed upside down on the coverslip and finally polymerized for 48h at 60°C in oven. The coverslips were removed by help of liquid nitrogen, and ultrathin sections of 50 nm were cut on a Leica Ultracut (Leica Mikrosysteme GmbH, Vienna, Austria) and picked up on a copper slot grid 2x1mm (EMS, Hatfield, PA, US) coated with a PEI film (Sigma, St Louis, MO, US). Sections were poststained with uranyl acetate (Sigma, St Louis, MO, US) 2% in H_2_O for 10 min, rinsed several times with H_2_O followed by Reynolds lead citrate in H_2_O (Sigma, St Louis, MO, US) for 10 min and rinsed several times with H_2_O.

### Mitochondria and mitochondria-endoplasmic reticulum contact analysis of EM images

Mitochondrial number, area, circularity, and perimeter were measured using Fiji/ImageJ. with adaptations to the methods described by Lam et al (https://www.ncbi.nlm.nih.gov/pmc/articles/PMC8465115/pdf/cells-10-02177.pdf) in si-CTRL and si-RyR1 myotubes from two independent cultures.

Only mitochondria that were entirely in the imaged field were analyzed.

Briefly, the outer mitochondrial membrane was outlined using the freehand selection tool. Each shape was added to the ROI manager, and the “analyze” menu was used to select the parameters to analyze. The measurements for each shape were obtained by clicking on “measure”. To analyze mitochondria-ER contacts, after identifying a mitochondrion in close contact with the ER membrane, we used the freehand line tool to draw a line spanning the length of the contact site. Each measurement was added to the ROI Manager and the “measure” function was used to determine the length of the mitocondria-ER contact. Similarly freehand line tool was used to draw a line between the two organelles. Each measurement was added to the ROI Manager and the “measure” function was used to determine the mitochondria-ER distance. Fifty images per biological replicate were acquired and 10 randomly selected images per sample were analyzed.

### Multiple pathway targeted lipidomic analysis

#### Sample preparation (complex lipid extraction)

Cell lysates were extracted by the addition of 200 μL of isopropanol (IPA)^1^. Samples were sonicated and centrifuged for 15 min at 4’000 g at 20°C (Hermle, Gosheim, Germany). Resulting supernatant was injected for LC-MS analysis. Then 100 μL of supernatant were taken and transferred to a LC-MS vial containing the internal standard (IS) mixture (75 isotopically labeled lipid species with multiple representatives of each lipid class with respect to varied fatty acid composition).

##### Internal standard mixture composition

Ultimate Splash One® mixture comprising 69 internal standards was purchased from Avanti (Alabaster, AL, USA). The additional isotopically labeled standards: HCER(d18:1/15:0)d7, LCER((d18:1/15:0)d7, DCER(d18:1/13:0)d7, arachidonic acid(20:4)d8, docosahexaenoic acid (22:6)d5 and eicosapentaenoic acid(20:5)d5, were purchased from Sigma-Aldrich.

#### Data acquisition -LC-MS/MS analysis

##### Next generation ‘omic-scale targeted lipidome analysis

Cell extracts were analyzed by Hydrophilic Interaction Liquid Chromatography coupled to electrospray ionization tandem mass spectrometry (HILIC ESI - MS/MS)^19^ in both positive and negative ionization modes^20^ using a TSQ Altis LC-MS/MS system (Thermo Scientific) in a dual-column setup. In both modes the chromatographic separation was carried out on an Acquity BEH Amide Premier, 1.7 μm, 100 mm × 2.1 mm I.D. column (Waters, Massachusetts, US). Mobile phase was composed of A = 10 mM ammonium acetate in Acetonitrile:H_2_O (95:5) and B = 10 mM ammonium acetate in Acetonitrile:H_2_O (50:50). The linear gradient elution from 0.1% to 20% B was applied for 2 min, then from 20% to 80% B for 3 min, followed by 3 minutes of re-equilibration to the initial chromatographic conditions. The flow rate was 600 μL/min, column temperature 45 °C and sample injection volume 2μl. Optimized HESI source parameters were set as follows: spray voltage 3500 V in positive mode and -2500 V in negative mode, sheath gas 60 au, aux gas 15 au, sweep gas 1 au, heated temperature and vaporizer temperature 350°C. Nitrogen was used as nebulizer gas and Argon as collision gas. Optimized compound-dependent parameters were used for data acquisition in timed selected reaction monitoring (tSRM) mode. Following the initial screening using six tSRM methods (englobing 1900 lipid species), robustly detected lipid species (using CV (peak area, height) < 30% across all QC samples and intensity cut-off = 1000 ion counts) were transferred for quantitative analysis. In addition, pooled QC samples (representative of the entire sample set) were analyzed periodically throughout the analytical run to correct for signal intensity drift (inherent to MS)^21^.

#### Data processing and quality assessment

Raw LC-MS/MS data was processed using the Trace Finder Software (version 4.1, Thermo Fisher Scientific The lipid abundance is reported as estimated concentration using a single-point calibration with a mixture of internal standards (IS) spiked at known concentration and designed to correct for diversity in fatty acid chain length and degree of unsaturation^22^. For each lipid species the concentration was calculated as follows: [Area of the analyte SRM]/[Area of the most structurally similar IS SRM] x [IS concentration in nmol/L]. The obtained tables (containing concentrations of detected lipids across all samples) were exported to “R” software http://cran.r-project.org/ for exploratory statistical analysis.

#### Data visualization

The pattern of variation and concentration distribution of each lipid species in the analyzed samples is presented by Box and Whiskers plots. Box and whisker plots display the full range of variation (whiskers: median with minimum maximum; boxes: interquartile range). Y-axis represents the lipid concentration (nmol/g of proteins) and x-axis represents the sample group.

### Chemicals

Fluo-4 AM, DMEM, CM-H2DCFDA, Rhod-2 AM, FBS, HS, NEAA, streptomycin-penicillin lipofectamine 3000, OptiMEM solutions were purchased from Life Technologies (Life Technologies, Thermo Fischer, Basel, Switzerland). Details on chemicals are provided in the supplementary information.

### Statistical analyses

Except for proteomic and metabolomic analyses, data are presented as mean ± SD. Statistical significance was determined using Student’s two-tailed t-tests to compare two groups. One-Way ANOVA was followed by Dunnett’s multiple comparisons post hoc tests to compare different groups when the main effect was significant. Data were analyzed using GraphPad Prism version 8.4.2. The level of significance was fixed at p ≤ 0.05.

## Results

### RyR1 transcripts is decreased in various human myopathies

To evaluate the role of RyR1 content in myopathies, using publicly available datasets we first measured the transcripts abundance of *RYR1* in skeletal muscle of patients suffering from various myopathies compared to healthy controls. We found an almost systematic significative decrease in *RYR1* transcripts in various forms of muscle diseases: 1) IM, necrotizing myopathy, IBM and polymyositis (Fig. 1A) (GSE39454 dataset ^12^), juvenile dermatomyositis (JDM) (Fig. 1B) (E-GEOD-3307 dataset^11^) and 2) muscular dystrophies such as DM2 (Fig. 1C) (GSE37084 dataset ^13^), presymptomatic DMD (Fig. 1D) (GSE6011 dataset^14^), LGMD2A, as well as Becker and symptomatic DMD (Fig. 1E) (E-GEOD-3307 dataset^11^).

**Fig. 1.**
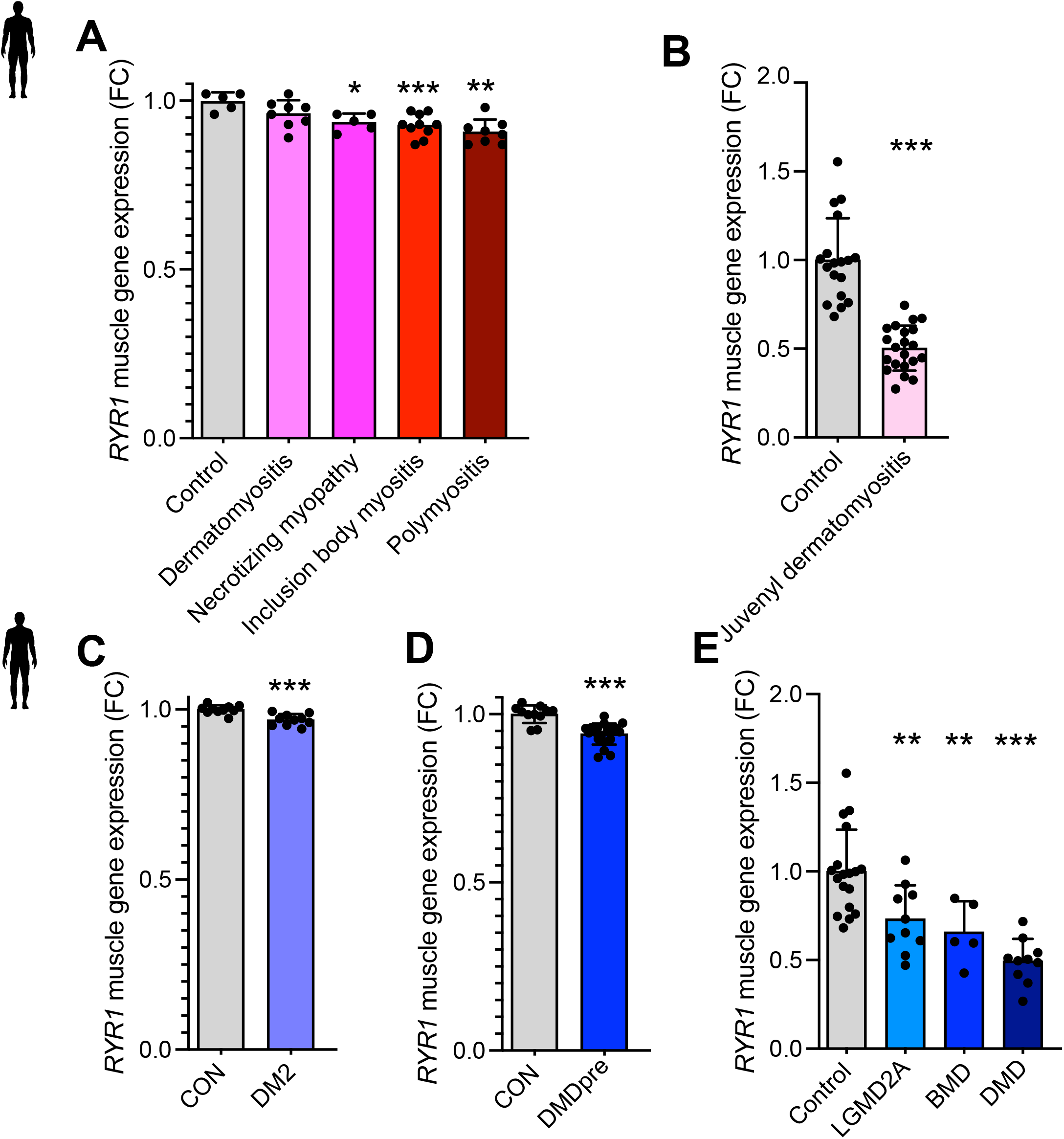
*RYR1* transcript abundance is decreased during human myopathies. RyR1 transcript abundance in muscle from patients affected with (**A**) dermatomyositis (n = 8), necrotizing myopathy (n = 5), inclusion body myositis (n = 10) and polymyositis (n = 8) compared to healthy controls (n = 5) (GSE39454 dataset), one-Way ANOVA followed by Dunnett’s multiple comparisons test. (**B**) juvenile dermatomyositis (n = 21) relative to healthy controls (n = 18) (E-GEOD-3307 dataset), Student’s two-tailed t-test. (**C**) Skeletal muscle *RYR1* expression in myotonic dystrophy 2 (DM2) (n = 10) relative to healthy controls (n = 10), Student two-tailed t-test. (**D**) Skeletal muscle *RYR1* expression in presymptomatic Duchenne Muscular Dystrophy (DMD) patients (n = 23) compared to healthy controls (n=14) (GSE6011 dataset), Student’s two-tailed t-test. (**E**) Skeletal muscle *RYR1* expression in Limb-girdle muscular dystrophy type 2A (LGMD2A) (n = 10), Becker muscular Dystrophy (BMD) (n = 5), DMD (n = 10) compared to relative healthy controls (n = 18) (E-GEOD-3307 dataset), one-Way ANOVA followed by Dunnett’s multiple comparisons test. All data are shown as mean ± SD. *P ≤ 0.05, **P ≤ 0.01, and ***P ≤ 0.001.

These data highlight a previously unreported common alteration of *RYR1* content in human muscular diseases.

### RyR1 protein is decreased in human non-*RYR1*-related myopathies

To confirm whether RyR1 protein content is also decreased in non-*RYR1*-RM, we analyzed a repertoire of human myopathies biopsies that we have created at the University Hospital of Lausanne and measured RyR1 protein levels by western blot. The target was to analyze muscle samples specifically from patients suffereing from welldefined myopathies with no link to RYR1-RM. We then selected inflammatory and mitochondrial myopathies groups. We observed a significant decrease in RyR1 protein levels, related to the muscle-specific myosin in IM muscle samples (IBM and dermatomyositis included) compared to age-matched control muscle biopsies (Fig. 2, A and B). We also observed the same significant decrease in RyR1 protein levels in samples from mitochondrial myopathy (MM) (Fig. 2, A and B). RyR1 protein is localized at the endo/sarcoplasmic reticulum where the sarco/endoplasmic ATPase channels (SERCA) play a central role in Ca^2+^ re-uptake into the ER/SR. Compared to the control healthy subjects, our investigations showed no significant alterations of the isoform 2 of SERCA channels (SERCA-2, Fig. 2, A and C), but a significant decrease in SERCA 1 isoform in both IM and MM samples (Fig. 2, A and D). Finally, no significant alterations were observed for the RyR1 channel stabilizer, FKBP12 (Fig. 2, A and E).

**Fig. 2.**
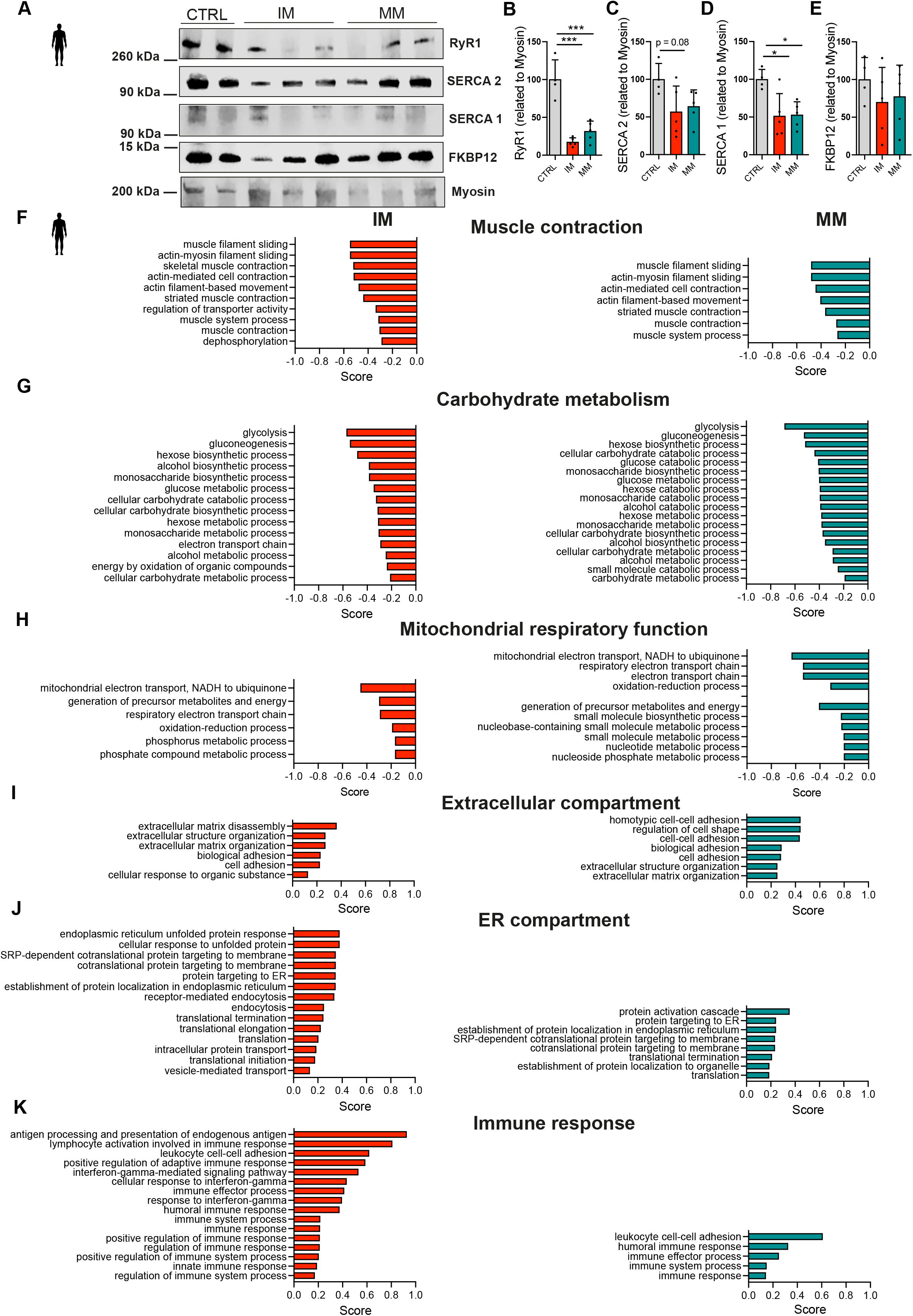
Non-*RYR1*-related myopathies show a decreased RyR1 protein content and similar altered pathways. (**A**) Western blot analysis of RyR1, SERCA 1, SERCA 2, FKBP12 and myosin (MF20) proteins in IM and MM compared to healthy controls and (**B-E**) quantification of RyR1, SERCA1, SERCA2 and FKBP12 protein levels in (**A**) from at least two different blots related to myosin (MF20) (n = 4 CTRL, 5 IM and 5 MM). Mean ± SD. One-Way ANOVA followed by Dunnett’s multiple comparisons test. (**F-H**) Proteomic analysis of protein groups related to Gene ontology Biological Processes (GoBP) that are significantly increased in IM (n = 4) and MM (n = 4) as compared to healthy controls (n = 4). (**I-K**) Proteomic analysis of protein groups related to Gene ontology Biological Processes (GoBP) that are significantly decreased in IM (n = 4) and MM (n = 4) as compared to healthy controls (n = 4) in IM and MM as compared to healthy controls. Benjamini-Hochberg corrected T-test. *P ≤ 0.05, ***P ≤ 0.001.

Both IM and MM muscle biopsies showed increased alpha-tubulin protein (Fig. S1, A and B), suggesting a cytoskeleton disorganization. We aimed at deciphering the molecular pathways altered in the non-*RYR1*-RM muscle samples showing decreased RyR1 protein content and thus performed a proteomic analysis. Compared to age-matched control samples, our results mostly showed similar alterations in IM and MM myopathies: 1) decrease of proteins involved in muscle contraction, carbohydrate metabolism and mitochondrial function (Fig. 2, F-H) and 2) increased levels of proteins found in the extracellular and ER compartments (Fig. 2, I-J). We also identified some specificities: IM samples showed a large immune system activation pattern, which was milder in MM (Fig. 1K), while the metabolic pathways were more altered in MM (Fig. 1H), supporting the characteristics of each myopathy. Proteomics studies have recently shown promise in characterizing immune complexes in IM ^23^ and our results again confirm the usefulness of this approach to tackle the immune alterations linked to IM.

Taken together, our results showed that two non-*RYR1*-RM, IM and MM, have decreased RyR1 protein levels, and share similarities in the alterations of muscle contraction, extracellular, ER and metabolic pathways.

### *In vitro* RyR1 protein depletion recapitulates the proteome changes observed in human

Myopathies are associated with various changes rending difficult to understand the molecular mechanisms underlying the disorders. Having established the pathways altered in IM and MM, both showing decreased RyR1 protein content, we then investigated if exclusive RyR1 protein depletion in muscle cells could lead to similar alterations in molecular pathways. We used the mouse muscle cell line C2C12, and the silencer select siRNA approach to deplete RyR1 protein in myoblasts. Myoblast differentiation was triggered the day after the transfection. RyR1 down-regulation in myoblasts leads to highly heterogenous, non-aligned or disorganized myotubes, completely different from the classic muscle cell phenotype (Fig. S2A). Quantitative polymerase chain reaction (q-PCR) showed depletion of *RYR1* transcripts at day 4 of transfection and day 3 of differentiation (D4TD3D) and D8TD7D (Fig. S2, B and I). Interestingly, we tested different doses of siRNAs and observed that the smallest applied dose (10 nM) was already effective to deplete RyR1 protein levels at D4TD3D (Fig. 3A). This effect was maintained until D7TD6D (Fig. 3B). RyR1 depletion in myoblasts was associated with a transient increase in inositol-1,4,5-triphosphate receptor (IP3R) protein content at D4TD3T that was normalized at D7TD6D (Fig. 3, A and B). High (50 nM and 100 nM) but not small (10 nM) doses of siRNAs were associated with increased SERCA1 protein (Fig. 3, A and B). However, all doses of siRNAs induced increased expression of alpha-tubulin as observed in human IM and MM muscles, and increased acetyl-tubulin, the stabilized tubulin form (Fig. 3, A and B).

**Fig. 3.**
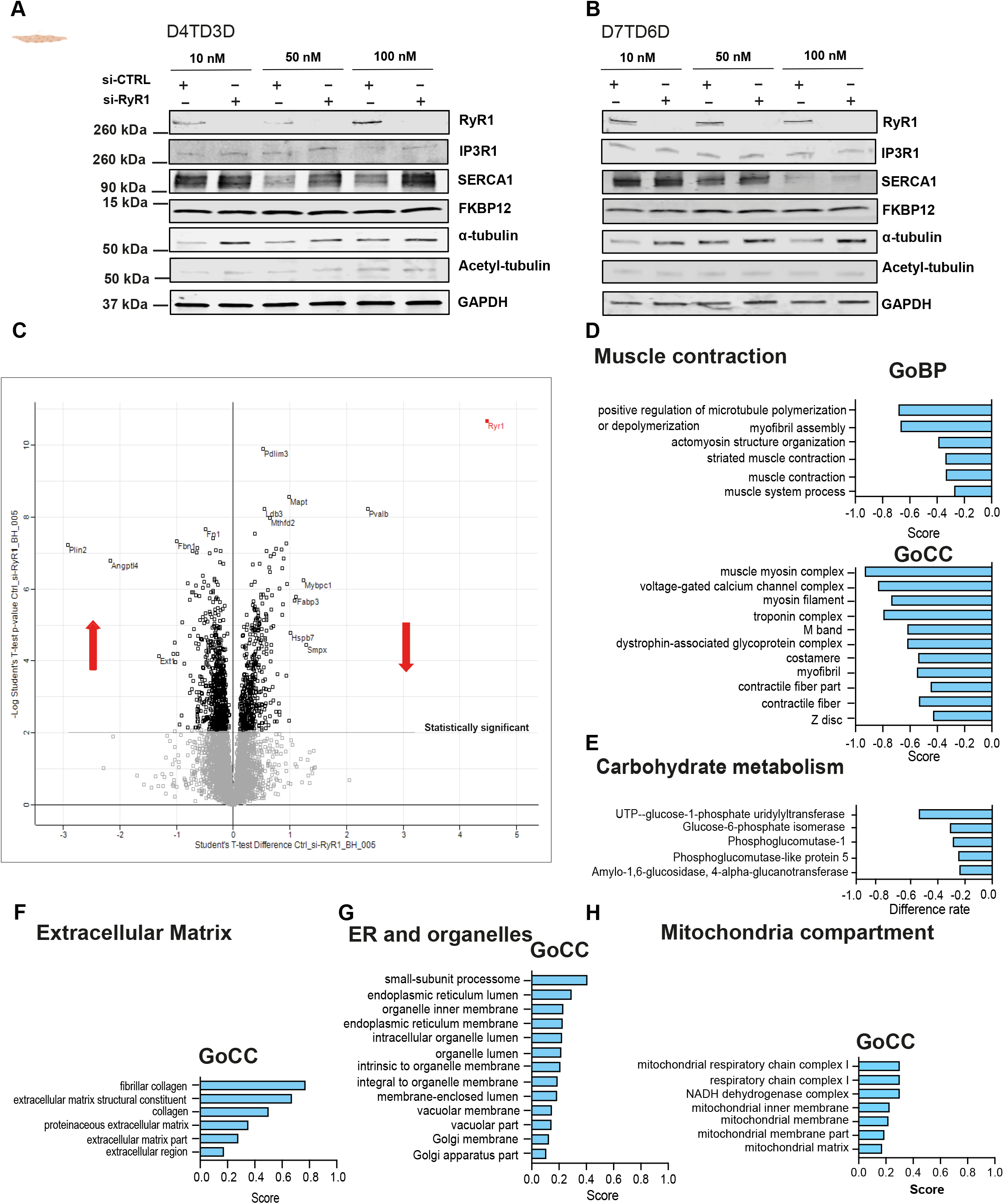
RyR1 depletion in myoblasts leads to myotubes with similar altered pathways as observed in human myopathies. Immunoblots of RyR1, IP3R1, SERCA, FKBP12, alpha tubulin, acetylated tubulin and GAPDH at (**A**) day 4 of transfection and 3 of differentiation (D4TD3D) and (**B**) day 7 of transfection day 6 of differentiation (D7TD6D). (**C**) Volcano plot of proteomic analysis showing the proteins significantly altered in si-RyR1 compared to si-CTRL myotubes. (**D and E**) Proteomic analysis of protein groups related to Gene ontology Biological Processes (GoBP) and cellular Component (GoCC) that are significantly downregulated in si-RyR1 as compared to si-CTRL myotubes. (**F-H**) Proteomic analysis of protein groups related to Gene ontology Biological Processes (GoBP) and cellular Component (GoCC) that are significantly upregulated in si-RyR1 as compared to si-CTRL myotubes. Benjamini-Hochberg corrected t-test.

Myosin heavy chain transcripts measured by q-PCR were not modified at D4TD3D (Fig. S2, C-D) but were decreased at D8TD7D (Fig. S2, J-K), supporting the phenotypic alterations observed in the muscle cells. GLUT4 transcript levels were increased at D4TD3D and normalised at D8TD7T (Fig. S2, E and L) while genes involved in oxidative stress were not changed at D4TD3D (Fig. S2, F-H) but significantly decreased at D8TD7D (Fig. S2, M-O), suggesting alteration in the oxidative defence in si RyR1 cells.

To have an overview of the alterations induced by RyR1 protein decrease in muscle cells, without any confounding factors from other altered ER Ca^2+^ channels, we used the smallest dose of siRNAs (10 nM) and performed a proteomic analysis at D7TD6D, a condition in which the only ER Ca^2+^ channel to be decreased was the RyR1, without any changes on SERCA or IP3R protein levels. Our proteomics volcano plot shows more than 4’000 proteins identified, of which more than 1’000 were significantly modified (Fig. 3D). Interestingly, we observed that RyR1 knock-down recapitulated the main features observed in human IM and MM by proteomics: 1) decreased muscle contraction proteins (Fig.3 D) and energy metabolism proteins (Fig. 3E), and 2) increased extracellular and ER compartments (Fig. 3, F-G).

These results clearly indicate that exclusive *in vitro* RyR1 protein depletion recapitulates most of the altered molecular pathways in human non-*RYR1*-RM, a result which suggests a broader role of RyR1 in muscle health, well beyond recessive *RYR1*-RM.

### RyR1 protein depletion induces defects in ER-mitochondria contact and mitophagy, and results in mitochondria aggregation

Our proteomics data showed a particular increase in the mitochondrial compartment in si-RyR1 myotubes (Fig. 3H). A deeper analysis revealed increased levels of mitochondrial specific proteins, which form a cluster composed of mitochondrial respiratory complex I, mitochondrial respirasome and mitochondrial chain complex associated with increased mitochondrial protein Tomm20 (Fig. 4A). This prompted us to investigate mitochondrial content, distribution and function in si-RyR1 cells compared to their controls. Using two different siRNAs against RyR1, we first stained the mitochondrial outer membrane protein Tomm20 by immunofluorescence. While mitochondria were homogenously distributed in si-CTRL myotubes, they appeared aggregated in some regions of the si-RyR1 myotubes, suggesting altered mitochondrial distribution (Fig. 4B). The same cells were stained with Hoechst, showing well aligned nuclei in si-CTRL myotubes while they were clustered in the Tomm20-positive regions in si-RyR1 myotubes. Acetyl tubulin used to delimit the shape of the cells as well as to validate our western blot data showed heterogenous distribution in si-RyR1 myotubes as compared to si-CTRL myotubes, suggesting altered cytoskeleton organization in si-RyR1 myotubes (Fig. 4B).

**Fig. 4.**
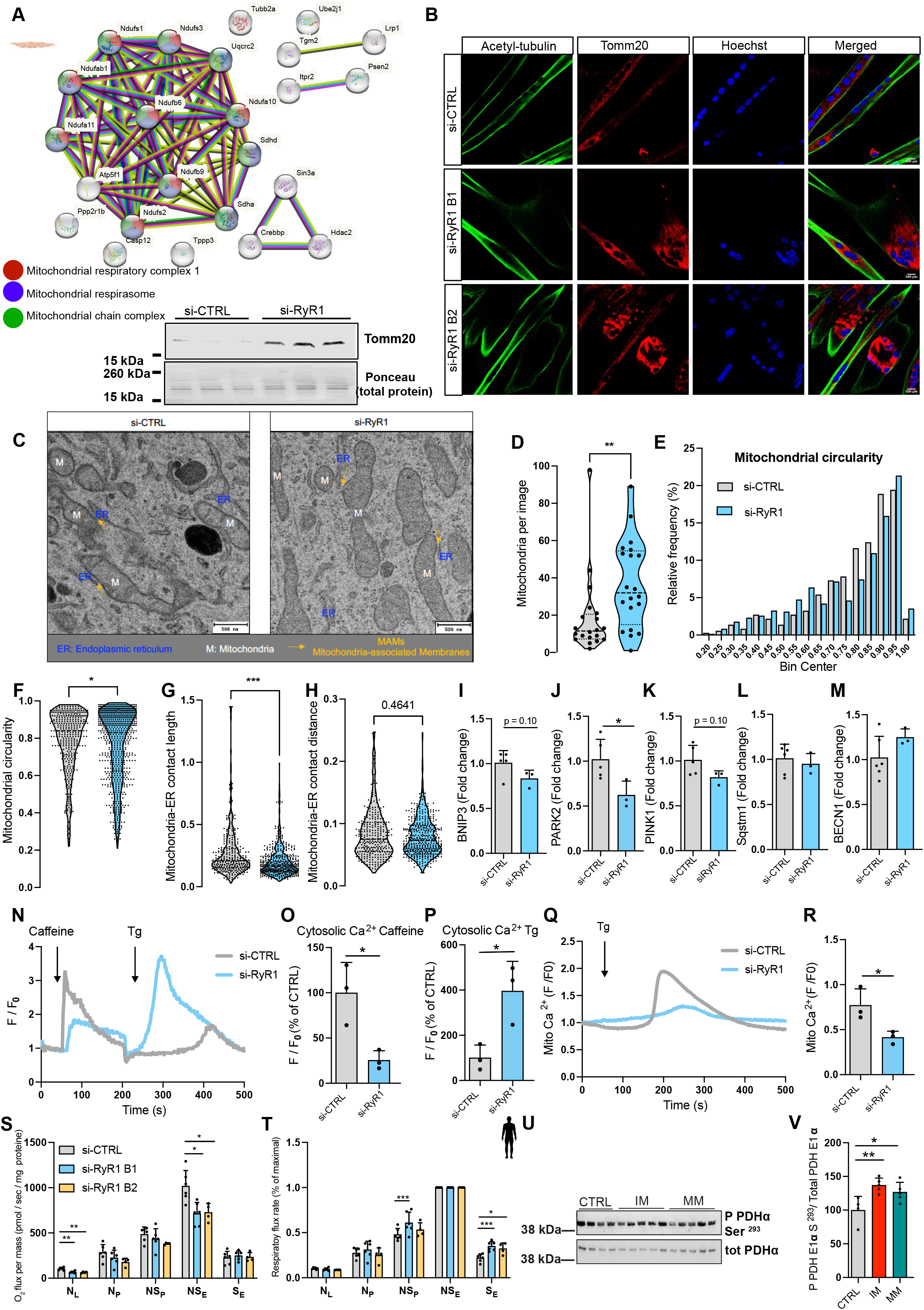
The reduction in RyR1 protein content impairs ER-mitochondrial contact, mitochondrial clearance, and function. (**A**) STRING protein-protein interaction network for mitochondrial proteins significantly increased in C2C12 si-RyR1 myotubes from the proteomics data and western blot showing increase of the mitochondrial outer membrane protein, Tomm20 in si-RyR1 myotubes at day 7 of transfection and 6 of differentiation compared to si-CTRL myotubes. (**B**) Representative confocal images of acetylated tubulin and Tomm20 immunofluorescence staining in C2C12 cells transfected with two different batches of si-RNAs as compared to si-CTRL at day 7 of transfection and 6 of differentiation. Hoechst was used to stain the nuclei. (n = three independent cultures). Magnification X40. (**C**) Representative transmission electron microscopy (TEM) images of si-CTRL and si-RyR1 myotubes at D7TD6D from two independent cultures. (**D**) Number of mitochondria counted per image. (**E**) Mitochondrial circularity frequency distribution (%) in si-CTRL and si-RyR1 myotubes. (**F**) Mitochondrial circularity in si-CTRL and si-RyR1 myotubes (371 contacts counted in siCTRL, 741 in siRyR1, n = two independent cultures). Student’s two-tailed t test. Mitochondria-ER length (**G**) and distance (**H**) (309 contacts counted in siCTRL, 457 in siRyR1, n = two independent cultures) Student’s two-tailed t test. (**I-M**) mRNA levels of genes involved in mitophagy (**I-K**) and autophagy (**L and M**) in si-CTRL and si-RyR1 myotubes (n = 6 and 3 independent biological experiments respectively). Student’s two-tailed t test. (**N**) Normalized Fluo-4 fluorescence traces in C2C12 si-CTRL and si-RyR1 myotubes and (**O and P**) quantifications of (**N**), (n = 3 independent biological experiments per group). Student’s two-tailed t test. (**Q**) Normalized Rhod-2 fluorescence traces in C2C12 si-CTRL and si-RyR1 myotubes and (R) quantifications of mitochondrial Ca^2+^ uptake in si-CTRL and si-RyR1 myotubes pre-treated with 10 μM dantrolene, (n = 3 independent biological experiments per group). Student’s two-tailed t test. (**S**) O_2_ flux per mass (pmol/sec/mg of protein) in si-CTRL and si-RyR1 myotubes at D7TD6D. MP_L_: malate pyruvate leak state. N_P_: N-linked OXPHOS state with ADP-stimulated. NS_P_: N- and S-OXPHOS pathways. NS_E_: ET state, noncoupled and S_E_, S-pathway; (n = 5 si-CTRL, 6 si-RyR1 batch 1 (B1) and 4 si-RyR1 batch 2 (B2) from independent biological experiments). One-way ANOVA with Dunnett’s multiple comparisons test. (**T**) Respiratory flux rate of si-CTRL and si-RyR1 from (S). (**U**) Immunoblots of phosphorylated PDH E1 α at serine 293 related to total PDH E1 α in human IM and MM (n = 4 CTRL, 5 IM and 5 MM) and (**V**) quantifications of (**U**) related to total protein. One-way ANOVA followed by Dunnett’s multiple comparisons tests. *p ≤ 0.05, **p ≤ 0.01, ***p ≤ 0.001.

As we observed a striking mitochondrial profile, we used transmission electron microscopy (TEM) to investigate mitochondria morphology and subcellular distribution in si-RyR1myotubes. While mitochondria were homogenously distributed in si-CTRL myotubes, we observed accumulated mitochondria with more elongated forms clustered in certain areas of si-RyR1 myotubes, with other regions almost devoid of mitochondria (Fig. 4C). TEM images were used to quantify mitochondrial number, mitochondrial morphology and mitochondrial ER contacts (according to ^24^). The number of mitochondria per field was increased in si-RyR1 myotubes compared to their controls (Fig. 4, C and D). In addition, we observed a significant decrease in mitochondrial circularity (Fig. 4 E and F). As shown in the frequency distribution graph (Fig. 4E) we observed a shift towards very elongated mitochondria (bin center between 0,25 and 0,60) in si-RyR1 myotubes, with a decrease of mitochondria with a circularity index between 0.65 and 0.9, also associated with some more perfectly round mitochondria. Together these results showed decreased mitochondria circularity and more elongated mitochondria in si-RyR1 compared to si-CTRL myotubes (Fig. 4E). These elongated mitochondria appear darker with unclear lumen, suggesting damaged mitochondria as observed during defect in mitophagy. Mitophagy is a quality control mechanism that ensures the elimination of damaged mitochondria ^25^ and ER contact sites with mitochondria form an exchange space called mitochondria-associated ER membranes (MAMs). The ER is responsible for the formation of mitophagosomes, where damaged mitochondria are ubiquitinated and dynamically encased ^26^. Impaired mitophagy triggers the accumulation of defective and non-functional mitochondria, which appear elongated ^27^. We then measured the ER contact sites with mitochondria in our cells. Mitochondria-ER contact length was reduced in si-RyR1 myotubes, without any alterations in the distance between the organelles (Fig. 4, G-H). To investigate whether the mitophagy process was affected in si-RyR1 myotubes, we measured the expression of mitophagy markers: PTEN-induced putative kinase 1 (PINK1), Parkinson protein 2 (PARK2), and BCL2 interacting protein 3 (BNIP3) by RT-qPCR. We observed a specific significant reduction of PARK2 expression (Fig. 4J) with no significant changes in BNIP3 and PINK1 transcripts levels (Fig. 4, I and K). Autophagy is a fundamental cellular process by which cells sequester intracellular constituents, including organelles and proteins, that are delivered to lysosomes for degradation and recycling of macromolecule precursors ^28^. As mitophagy is a specific form of autophagy, we also measured transcript levels of genes involved in the autophagy process. No significant mRNA alterations were observed for the classical autophagy genes sequestosome 1, SQSTM1 and BECLIN1, BECN1 (Fig. 4, L and M). PARK2 encodes for Parkin, which is an important protein involved in the mitophagy process as it promotes the recruitment of p62, a ubiquitin-binding adaptor protein responsible of the recruitment of ubiquinated cargo in autophagosomes by binding to LC3. Accordingly, in mouse embryonic fibroblasts, depletion of Parkin has been shown to completely impair mitophagy ^26^. We have recently reported a specific defect in mitophagy in DMD, with decrease in PINK1, PARK2 and BNIP3 mRNAs. Interestingly, Urolithin A treatment to restore mitophagy alleviates most signs of the disease^29^.

Together, our results show that RyR1 protein decrease impairs ER-mitochondria tethering and mitochondria accumulation potentially through defects in mitophagy.

### RyR1 protein depletion alters ER-mitochondria Ca^2+^ transfer and mitochondrial function

ER contact sites with mitochondria at MAMs also allow a direct transfer of Ca^2+^ from the ER to the mitochondria by connecting the voltage-dependent anionic channel (VDAC) on the outer mitochondrial membrane to the IP3R on the ER membrane ^30^. Alterations to this process disturb ER Ca^2+^ homeostasis ^31^. We investigated whether the decrease in ER-mitochondria contact length in si-RyR1 may affect Ca^2+^ transfer from the ER to the mitochondria. We first investigated the capacity of our cells to release Ca^2+^ from the ER. Using the cytosolic Ca^2+^ probe Fluo4 and 2.5 mM caffeine stimulation to open the RyR1, we observed that ER Ca^2+^ release observed in si-CTRL myotubes was blunted in si-RyR1 cells (Fig. 4, N and O). This result was expected due to the depletion of RyR1 protein in si-RyR1 cells. To ensure that si-RyR1 cells still contain Ca^2+^ within the ER, we stimulated the same cells being exposed to caffeine with 1 μM of thapsigargin, which inhibits Ca^2+^ uptake through the SERCA pumps leading to Ca^2+^ leak through IP3R. Interestingly, si-RyR1 cells elicited nice Ca^2+^ transients that were much lower in si-CTRL cells whose ER were previously depleted (Fig. 4, N and P). These results suggest that si-RyR1 protein depletion does not significantly alter ER Ca^2+^ load as compared to si-CTRL. To then investigate Ca^2+^ transfer from the ER to the mitochondria, we loaded the cells with the mitochondria Ca^2+^ probe Rhod-2/AM and stimulated them with 1 μM of thapsigargin in the presence of dantrolene (a known inhibitor of the RyR1 channel) to avoid any Ca^2+^ input from RyR1 channels in si-CTRL cells. Our results clearly showed increased mitochondrial Ca^2+^ uptake in si-CTRL cells, which was blunted in si-RyR1 myotubes (Fig. 4, Q and R). These results indicate that the impaired ER-mitochondria contact length alters Ca^2+^ transfer from the ER to the mitochondria in si-RyR1 myotubes.

We investigated whether the above described alteration may affect mitochondrial function by using high-resolution respirometry to measure mitochondrial respiration. We assessed leak respiration (MP_L_) with malate and pyruvate (at non-phosphorylating state) and observed a significant decrease in si-RyR1 compared to si-CTRL myotubes (Fig. 4S). OXPHOS-state (N_P_), measured by adding saturating concentrations of ADP followed by glutamate, was not significantly altered in si-RyR1 compared to si-CTRL myotubes (Fig. 4S). When oxygen consumption was further increased by the addition of succinate to induce convergent electron transfer into the Q junction (NADH- and succinate-linked pathways, NS_P_ state) we did not either observe a significant difference in si-RyR1 compared to si-CTRL myotubes (Fig. 4S). However, when we assessed maximal electron transfer system activity by the addition of the protonophore FCCP (NS_E_ state), the maximal respiration capacity was significantly decreased in si-RyR1 compared to si-CTRL myotubes (Fig. 4S). Inhibition of NADH-linked respiration by rotenone did not show a significant alteration in non-phosphorylated succinate-linked pathway (S_E_) in si-RyR1 compared to si-CTRL myotubes (Fig. 4S). These results were validated with a second si-RNA against RyR1 (Fig. 4S). We then calculated the respiration flux rate as a percentage of the maximal respiration capacity. This approach provides non-quantitative but qualitative contribution of each state to the maximal respiration ^32^. Our results showed a significant increase in the contribution of succinate-linked pathway to the maximal respiration especially in the non-phosphorylated state in si-RyR1 myotubes using both si-RNAs against RyR1 compared to their controls (Fig. 4T). These findings indicate a particular involvement of the succinate-linked pathway in mitochondrial respiration despite the overall decreased maximal respiration capacity in si-RyR1 compared to si-CTRL myotubes, suggesting a higher lipid oxidation in si-RyR1 compared to si-CTRL myotubes.

Frozen human muscle samples are not appropriate to investigate mitochondrial Ca^2+^ uptake and function as freezing damage mitochondrial membranes. We measured mitochondrial protein Tomm20 as well as OXPHOS proteins and observed no significant differences in Tomm20 related to total protein, or in CI, II, III and IV complex proteins levels related to CV ATPase, as compared to control muscles (Fig. S1, C-I).

We then indirectly measured the activation of pyruvate dehydrogenase (PDH), a key enzyme involved in the conversion of pyruvate to Acetyl-CoA by measuring its phosphorylation level by western blot and used it as a readout of mitochondrial Ca^2+^ uptake in human myopathy samples ^2^. PDH phosphorylation level on Ser^293^ (P PDH E1 Ser^293^) was significantly increased in both human IM and MM, suggesting a potentially decreased mitochondrial Ca^2+^ uptake and PDH activation (Fig. 4, U and V), which could contribute to impaired mitochondrial function in human muscle.

Together, our results show that RyR1 protein decrease, which alters ER mitochondrial tethering, also impairs mitochondrial Ca^2+^ uptake and mitochondrial function.

### RyR1 depleted myotubes show accumulation of sphingolipids, phospholipids and glycerolipids

Our previous data suggest a higher lipid oxidation in si-RyR1 compared to si-CTRL myotubes. Interestingly, our proteomics data showed an important cluster of proteins related to lipid metabolism, among which sphingolipid, glycerolipid and phospholipid biosynthetic and metabolic processes, in si-RyR1 myotubes compared to their controls (Fig. 5A). Detailed analyses reveal a series of lipid metabolism proteins, with the top modified protein being the lipid droplet (LD) coating protein perilipin 2 (Plin2) (Fig. 5B). The proteomic analysis clearly showed an inverse correlation between RyR1 and Plin2 expression (Fig. 5C). We confirmed by western blot the increased Plin2 levels in si-RyR1 compared to si-CTRL myotubes (Fig. 5, D and E). To investigate whether increased LD coating protein was associated with increased LDs, we used the BODIPY to stain LDs in our cells. Interestingly, using two different si-RNAs against RyR1, our results showed accumulation of LDs in si-RyR1 compared to si-CTRL myotubes (Fig. 5F). Together, these results clearly suggest altered lipid dynamics in si-RyR1 compared to si-CTRL myotubes. To identify which lipid species are modified in an unbiased way, we performed a semi-quantitative lipidomic analysis. Our results nicely show a large increase in the proportion of sphingolipids (Fig. 5G), phospholipids (Fig. 5H) and phosphoglycerols (Fig. 5I) in si-RyR1 compared to si-CTRL myotubes, supporting all our previous observations. In addition, we observed a large increase in ceramides and its precursors dihydroceramides, sphingomyelins as well as increased hexosyl-ceramides (Fig. 5G). The potential role of RyR1 in ceramide synthesis is novel and exciting as regard to the understanding of DMD pathogenesis, since our bioinformatic analysis reveals decreased *RYR1* transcripts in several myopathies including DMD, in which we have very recently reported the deleterious role of ceramide *de novo* synthesis in the pathogenesis of the disease ^33^. Indeed, the increased ceramide species in our si-RyR1 cells mostly correspond to the ceramide species altered in DMD ^33^ The most modified phospholipids are phosphatidylinositols and phosphatidylethalonamides (Fig. 5H). Among the neutral lipids, phosphoglycerols, diglycerides and fatty acids are the most upregulated with three monoglycerides that were downregulated (Fig. 5I). Together these results confirm alteration of lipid metabolism in response to RyR1 depletion.

**Fig. 5.**
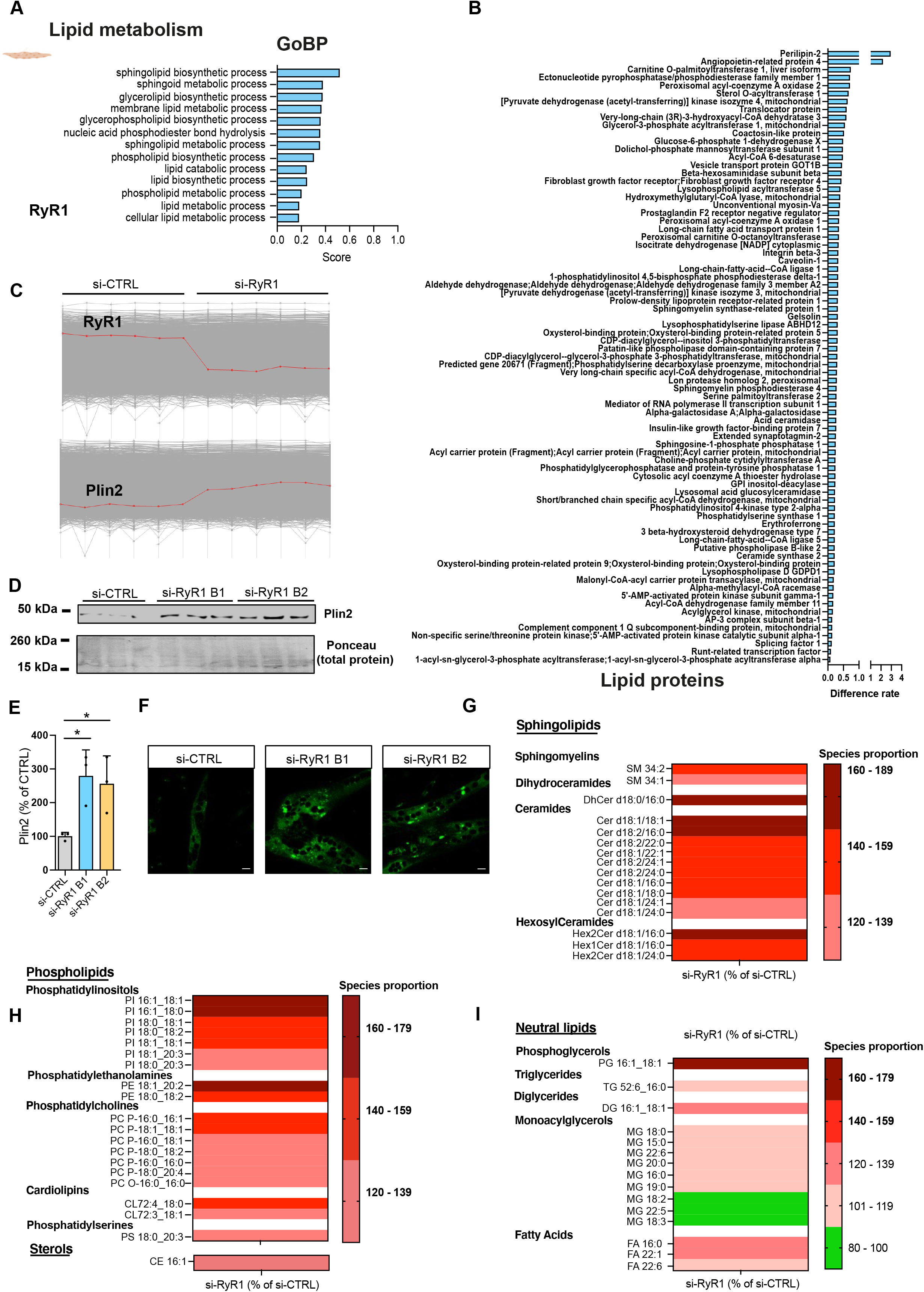
The reduction in RyR1 protein content alters lipid homeostasis in muscle cells. (**A**) Proteomic analysis showing protein groups related to Gene ontology Biological Processes (GoBP) involved in lipid metabolism that are significantly upregulated in si-RyR1 as compared to si-CTRL myotubes. Benjamini-Hochberg corrected T-test. (**B**) Detailed list of proteins involved in lipid homeostasis that are significantly upregulated in si-RyR1 as compared with si-CTRL myotubes. (**C**) Scheme of RyR1 and Periliplin 2 (Plin2) inverse association in si-CTRL and si-RyR1 proteomic samples. (**D**) Representative immunoblot of Plin2 protein in si-CTRL and si-RyR1 (**B1 and B2**) myotubes at D7TD6D and (**E**) quantifications of Plin2 in (**D**) related to total protein, (n = 3 independent biological experiments per group). One-Way ANOVA followed by Dunnett’s multiple comparisons tests. (**F**) Representative confocal images from three independent biological experiments of lipid droplet staining in si-CTRL and si-RyR1 myotubes at D7DD6T. Magnification X 40. Scale bar 500 μm. (**G-I**) Lipidomic analysis of si-CTRL and si-RyR1 myotubes at D7TD6D from two independent cultures. Lipid species were expressed as a proportion of total species detected. Only the lipid species that showed changes in both independent biological experiments were considered as altered. Data are the mean of the two independent biological experiments expressed as percentage of si-CTRL. *p ≤ 0.05.

Taken together, a RyR1 protein decrease is associated to altered lipid metabolism both in cells and in human myopathy.

### RyR1 protein decrease in muscle is associated with ER stress in cultured cells, mouse and human muscle samples

So far, our results indicate that the RyR1 protein decrease *in vitro* recapitulates most of the features of human non-*RYR1*-RM myopathies characterized by a decreased RyR1 protein content. Our results highlighted alterations in ER-mitochondria contact and Ca^2+^ transfer, mitophagy and mitochondrial function, as well as in lipid homeostasis. All those alterations are observed in myopathies but their causes are not elucidated. Thanks to our *in vitro* models we now prove a role of RyR1 protein decrease in those processes, a demonstration that could not be possible in human samples. However, it is unknown how the reduction in RyR1 protein mediates all those alterations. Decreased ER-contact sites and Ca^2+^ transfer from ER to the mitochondria has been observed in the mouse model of DMD, the *mdx* mouse, where it has been attributed to ER stress. Accordingly, *mdx* mice treatment with TUDCA, an ER stress inhibitor, restored Ca^2+^ transfer from the ER to the mitochondria and improve signs of the disease ^31^. Interestingly, we have shown defects in mitophagy in *mdx* mouse muscles, which we linked to a defect in ER-mitochondria tethering ^29^. Besides, ER is an important site of lipid metabolism since many enzymes involved in lipid metabolism are located at the ER ^34^. Thus, ER stress may alter lipid metabolism. Activation of ER stress has been shown as a central mechanism of the pathogenesis of obesity and peripheral insulin resistance ^35^ and increasing research evidence shows association of ER stress with myopathies, including IM and DMD^10,31,36^.

ER stress triggers a physiological response called the unfolded protein response (UPR), whose prolonged activation can lead to cell death. UPR activation is regulated by three distinct transmembrane proteins and their signalling pathways: the pancreatic ER kinase (PRK)-like ER kinase (PERK), the activating transcription factor 6 (ATF6), and the inositol-requiring enzyme 1 (IRE1α)^31^. In normal conditions, these transmembrane proteins are inactivated by the chaperone-binding protein/glucose regulated protein 78 (GRP78-Bip) ^37^. In response to ER stress, GRP78-Bip dissociates from the transmembrane proteins and binds misfolded proteins, leading to the activation of the UPR response. Accordingly, an increase in GRP78-Bip protein has been shown as a strong marker of ER stress ^38^. We then investigated key ER stress markers in si-RyR1 myotubes compared to their controls. Interestingly, our data showed increased GRP78-Bip in si-RyR1 myotubes compared to si-CTRL myotubes (Fig. 6, A and B). ER stress outcomes are activation of pro-survival factors such as activation through phosphorylation of the eukaryotic initiation factor 2 alpha (EIF2*α*), which promotes a pro-adaptive signaling pathway by the inhibition of global protein synthesis and selective translation of Activating Transcription Factor 4 (ATF4) ^39^. Our investigations showed increased phosphorylated forms of EIF2*α* but also of its non-phosphorylated forms, rending the ratio of phosphorylated EIF2*α* non modified in si-RyR1 myotubes compared to their controls (Fig.6, A and C). This result suggests that the ER stress response in si-RyR1 cells might be inefficient to promote cell survival. Indeed, during conditions of prolonged ER stress, pro-adaptive responses fail, and apoptotic cell death ensues by the pro-apoptotic protein CCAAT-enhancer-binding protein homologous protein CHOP-DDIT3 upon ATF4 induction. This leads to the induction of several pro-apoptotic genes and suppression of the synthesis of anti-apoptotic Bcl-2 proteins ^39^. Accordingly, CHOP-DDIT3 protein was increased in si-RyR1 muscle cells (Fig. 6, A and C). These results clearly show increased ER stress response and its apoptotic effector triggered by RyR1 protein absence. ER stress-mediated apoptosis has been implicated in the pathogenesis of several conformational diseases ^40^.

**Fig. 6.**
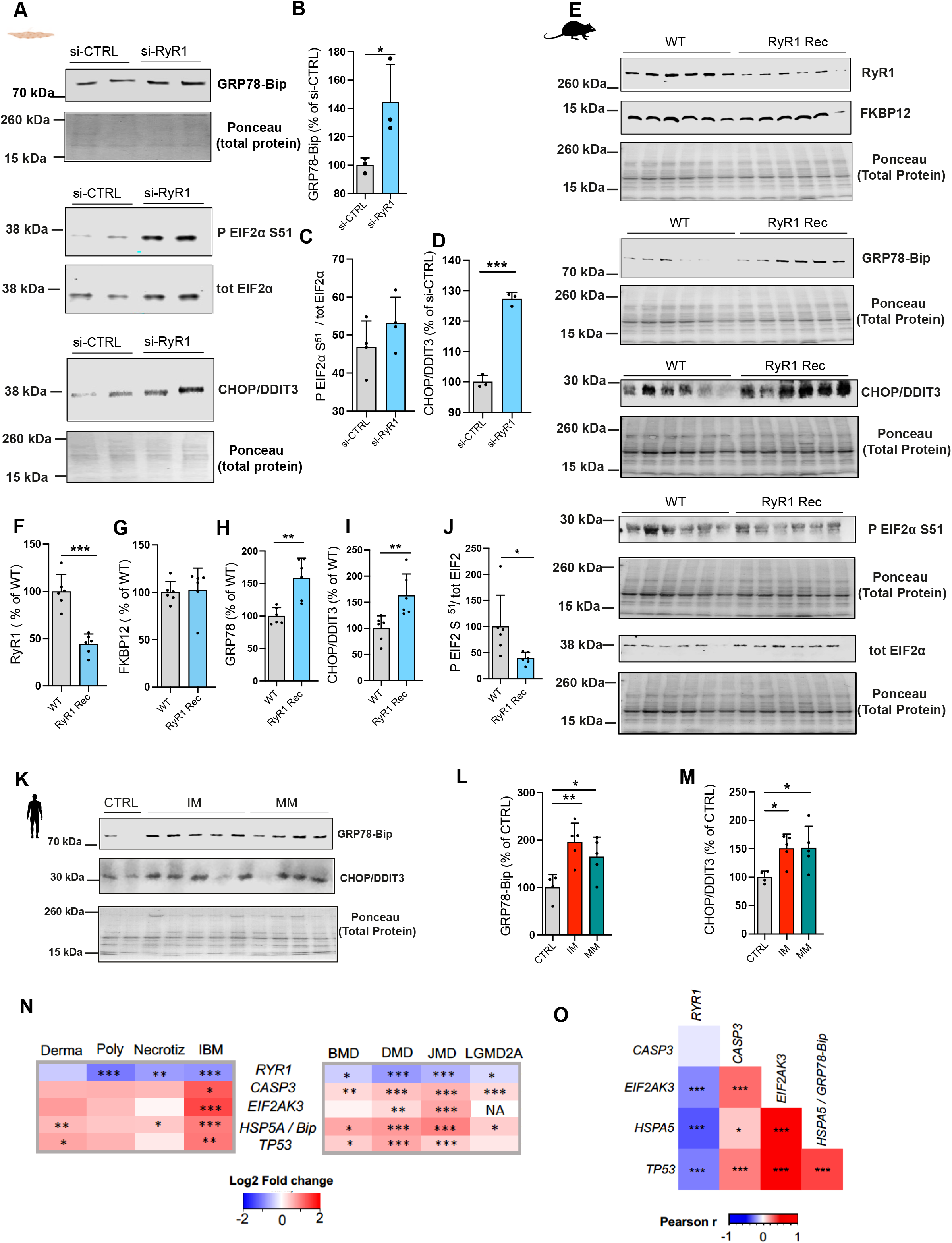
The decrease in RyR1 protein content induces ER stress in muscle cells. (**A**) Representative immunoblots of GRP78-Bip, CHOP-DDIT3, EIF phospho S51 and total EIF2 protein levels in C2C12 si-CTRL and si-RyR1 myotubes at D7TD6D and (**B-D**) quantification of proteins in (**A**) related to total protein, (n = 3-4 independent biological experiments). Student’s two-tailed t test. (**E**) Immunoblots of RyR1, FKBP12, GRP78-bip, CHOP-DDIT3, EIF2 α phospho S51, total EIF2 α protein levels in WT and RyR1 Rec mice sacrificed 75 days after tamoxifen injection. (**F**) Quantifications of RyR1 protein levels in (**E**) related to total protein. (**G**) Quantifications of FKBP12 protein levels in (**E**) related to total protein. (**H**) Quantifications of GRP78-Bip protein levels in (**E**) related to total protein. (**I**) Quantifications of CHOP-DDIT3 protein levels in (**E**) related to total protein. (**J**) Quantifications of EIF2 α phospho S51 related total EIF2 α in (E). (N = 6 mice per group). All the ER stress proteins were quantified from four independent blots, which are averaged. Student’s two-tailed t test. *p ≤ 0.05, **p ≤ 0.01, ***p ≤ 0.001. (K) Immunoblots of GRP78-Bip and CHOP-DDIT3 protein levels in human muscles biopsies from IM (n= 5) and MM (n=5) compared to control healthy subjects (n= 4). (**L**) Quantifications of GRP78-Bip protein levels in (**K**) related to total protein. (M) Quantifications of CHOP-DDIT3 protein levels in (**K**) related to total protein. All human proteins were quantified from at least two independent blots, which are averaged. One-Way ANOVA followed by Dunnett’s multiple comparisons tests, (**N**) heat map of RyR1 and ER stress transcripts in inflammatory myopathies and dystrophies respectively from a publicly available dataset (GSE39454). (**O**) Correlogram showing Pearson correlations between *RYR1* transcripts levels and ER stress markers in *postmortem* skeletal muscle biopsies from the Genotype-Tissue-Expression (GTEx) v7 dataset, (n =491), with Student p-values indicated *p ≤ 0.05, **p ≤ 0.01, ***p ≤ 0.001

To evaluate the relevance of our observations *in vivo*, we used *in vivo* inducible RyR1 recessive mouse muscles, which lead to half depletion of RyR1 protein and recapitulate most features of the Central Core Disease ^41^ to measure ER stress protein levels. Our results first confirmed a 50% decrease of RyR1 protein in RyR1 Rec muscles compared to their control wild type (WT), with no modifications on the RyR1 stabiliser protein FKBP12 (as in our cellular models) (Fig. 6, E-G). Interestingly, RyR1 Rec muscles showed increased GRP78-Bip and CHOP-DDIT3 protein levels as observed in our cellular models (Fig. 6, E and H and I) associated with decreased EIF2*α* phosphorylation levels (Fig. 6, E and J). These results confirm *in vivo* the role of RyR1 protein decrease in ER stress induction.

Next, we confirmed increase in GRP78-Bip and CHOP-DDIT3 in our human muscles from IM and MM (Fig. K-M). These results strongly suggest an association between decreased RyR1 protein levels and increased ER stress-mediated apoptosis also in humans.

We finally used bioinformatics tools to investigate the association between ER stress markers and RyR1 transcript levels by exploiting publicly available datasets. Our results interestingly showed an increase in ER stress markers in various human IM and dystrophies (IBM, JDM, LGDM2A, Becker and DMD), which correlates with the decreased *RYR1* transcript levels (Fig. 6, N-O).

Taken together, our data clearly show that RyR1 depletion induces ER stress in cells, mice and humans and that there is a negative association between *RYR1* transcript levels and ER stress markers in human muscle cell during myopathies.

## Discussion

Our investigations reveal a previously unreported significant decrease of *RYR1* transcripts in various myopathies, that may suggest a potential role of RyR1 in their pathogenesis. All the myopathies screened in this paper have well-established causes and thus a systematic decrease in *RYR1* transcripts either reflects muscle remodeling or highlights a potential key role of RyR1 in the pathophysiology of muscle disease. This common decrease in *RYR1* transcripts in myopathies might constitute a new diagnosis tool of muscle diseases. The unsolved question is how *RYR1* transcripts are affected during myopathies, a question that future investigations should help clarifying.

We observed decreased RyR1 protein content in IM, of which most forms are associated with decreased *RYR1* transcripts, and in MM, another non-RyR1 myopathy. Decreased RyR1 protein content in muscle samples from IBM has been previously reported (Amici, 2017) and the authors suggested a role of calpain-activated protein degradation in the process. We here confirm in two different IM forms (dermatomyositis and IBM) a decreased RyR1 protein content. While proteolysis might play a role in the decreased RyR1 protein content in IM muscle, it might not be the only mechanism involved in the process. Indeed, the decrease in *RYR1* transcripts could not only account for muscle disuse as it has been also observed at earlier and pre symptomatic stages of DMD. We have previously reported sphingolipid accumulation in pre symptomatic DMD muscle samples^33^, even though sphingolipid biosynthetic pathways correlate with the severity of the disease ^33^. We also recently provided evidence of the role of the deleterious role of sphingolipid accumulation in the development of sarcopenia (https://doi.org/10.1038/s43587-022-00309-6). Our study here clearly shows a sphingolipids accumulation in response to RyR1 protein depletion.

We hypothesized in this study that RyR1 protein depletion may induce ER stress to alter lipid metabolism, ER-mitochondria Ca^2+^ transfer, mitophagy and mitochondrial dysfunction. But it is also well established that accumulated misfolded proteins, excess numbers of damaged lipids, or a Ca^2+^ imbalance can also induce ER stress ^31,42^. ER stress may thus potentially constitute a cause but also a consequence of the alterations induced by the RyR1 protein depletion. Despite the impairment of ER-mitochondria Ca^2+^ transfer in si-RyR1 myotubes, our investigations did not reveal significant alteration of ER Ca^2+^ content in si-RyR1 myotubes compared to their controls that might trigger ER stress. However, the ER is not only the site of protein folding but also an important site of lipid metabolism since many enzymes involved in lipid metabolism are located at the ER ^34^. RyR1 being localized at the ER, it may play a direct role in lipid synthesis/metabolism and/or modulation of the ER structure and function for protein folding. The absence of RyR1 may thus induce altered lipid dynamics and/or protein folding, which finally will lead to ER stress. Further investigations are necessary to elucidate the role of ER stress in RyR1 protein decrease-induced muscular disorders. This might open rapidly implementable therapeutic approaches to alleviate the muscular disorders induced by decreased RyR1 content, as ER stress inhibitors are already used in clinics.

ER stress is more and more associated to clinical conditions including myopathies. A recent study has reported ER stress in a mouse model of I4898T RyR1 mutation, the most studied mutation on the RyR1 gene, with an UPR response essentially mediated by CHOP ^36^. We report for the first time an increase in GRP78-Bip and CHOP in si-RyR1 myotubes, in human IM and MM muscles, as well as in inducible half recessive RyR1 mouse muscles, indicating activation of ER stress and its pro-apoptotic effector. Moreover, our bioinformatic data brings evidence of a negative association between ER stress markers and *RYR1* muscle content in various myopathies. Of note a kinase activating the pro-survival factor EIF2, EIF2AK3, are also increased in human myopathies and negatively correlate with RyR1 levels. EIF2A kinases have been shown to increase in response to decreased EIF2 phosphorylation as an attempt to rescue cell survival. However, this adaptive mechanism is rarely successful. Indeed, declined EIF2 phosphorylation has been shown to be associated with increased CHOP and EIF2 kinases protein levels, but not with their activity, in aged muscles ^43^. Accordingly, in some cases, deletion of the gene encoding CHOP preserves tissue function in case of chronic ER stress/maladaptive UPR^44^.

A previous study has investigated Ca^2+^ dynamics in response to RyR1 protein decrease in mouse C2C12 muscle cells^45^. However, all investigations were made in conditions where RyR1 depletion was associated with increased IP3R protein levels and accordingly, the authors have observed increased ER-mitochondria Ca^2+^ transfer in si-RyR1 cells. We also observed IP3R protein upregulation at the earliest stages of RyR1 depletion in muscle cells. To unravel the specific role of RyR1 protein content in muscular disorders, we tried to avoid confounding factors from other Ca^2+^ channels located at the ER and thus performed all our investigations at a time point where IP3R and SERCA proteins were unmodified in si-RyR1 cells compared to their controls. Indeed, myopathies are associated with a myriad of protein changes rending difficult the elucidation of the causes and the consequences of the disorders.

It has been previously reported an increase in all LC3 forms in inducible half recessive RyR1 depletion, suggesting defects in autophagy ^41^. All forms of LC3 were also accumulated in si-RyR1 cells (data not shown). However, LC3, even if well-known as an autophagosome marker can also be incorporated into protein aggregates independently of autophagy, imposing caution in the interpretation of LC3 I and II simultaneous increase. We here report a defect in mitophagy in si-RyR1 cells compared to their controls (which does not exclude a global defect in autophagy), supported by decreased ER-mitochondria contacts, decreased PARK2 levels and accumulation of long and non-functional mitochondria. As PARK2 promotes the recruitment of p62, responsible of the recruitment of ubiquinated cargo in autophagosomes by binding to LC3, we cannot exclude that PARK2 decrease might impair p62 recruitment and LC3 binding and use leading to their accumulation. Interestingly, mitochondrial dysfunction has also been linked to core formation in *RYR1*-RM, which show focal loss of mitochondria ^46^.

## Conclusions

Our study reveals broad alterations in RyR1 content in myopathies beyond the *RYR1*-RMs. Using a translational approach, we uncover the important molecular pathways altered by decreased RyR1 content. RyR1 depletion in myotubes, with no modifications on other channels located on the ER, phenocopies most of the biological pathways alterations onbserved in two different human non-*RYR1*-RM (IM and MM), suggesting similarities in the underlying mechanisms. Our *in vitro* models help deciphering the molecular pathways altered in response to RyR1 protein decrease. We identified decreased ER-mitochondria contact and decreased mitophagy gene PARK2 associated to mitochondria aggregation and dysfunction in absence of RyR1 protein. Moreover, RyR1 depletion alters lipid homeostasis leading to accumulation of deleterious forms of sphingolipids. All these alterations are observed in myopathies. Importantly, RyR1 protein decrease is associated with ER stress, which is more and more associated with myopathies, in cells, mouse and human muscles, pointing to an important role of RyR1 protein decrease and ER stress in the pathogenesis of myopathies.

## Data Availability

All data produced in the present work are contained in the manuscript

## Figures legends

**Fig. S1.**
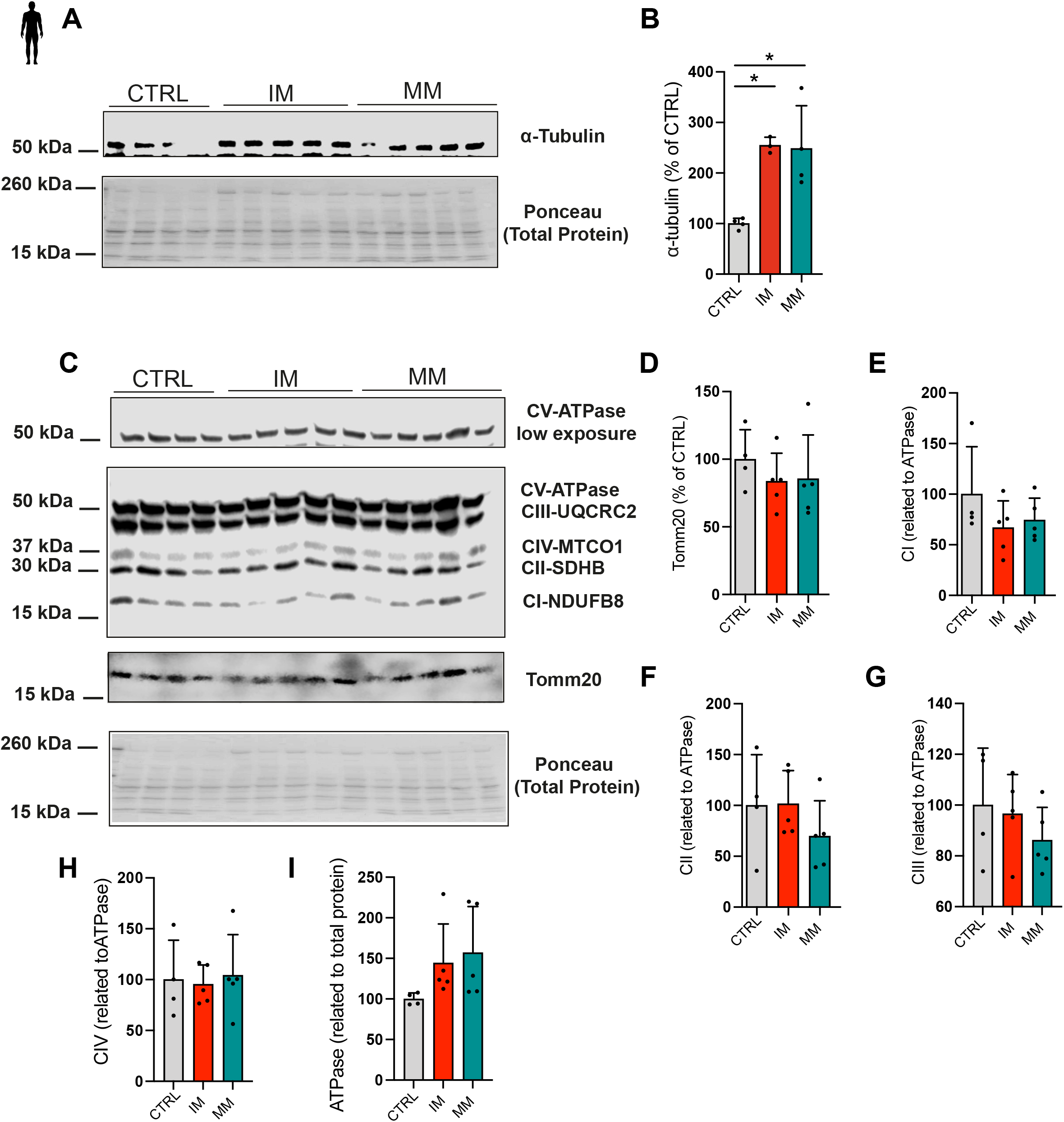
Cytoskeleton and mitochondrial proteins in muscle samples from IM and MM patients. (**A**) Immunoblots of alpha-tubulin and (**B**) its quantifications related to total protein in muscle samples from inflammatory and mitochondrial myopathies (n= 4 controls, 5 IM et 5 MM). (**C**) Immunoblots of OXPHOS and Tomm20 proteins in muscle samples from inflammatory and mitochondrial myopathies and (**D-I**) their quantifications (n= 4 controls, 5 IM et 5 MM). OXPHOS proteins CI, CII, CIII and CIV were reported to CV ATPase and CV ATPase levels were related to total proteins. One-way ANOVA with Dunnett’s multiple comparisons test. *p ≤ 0.05, **p ≤ 0.01, ***p ≤ 0.001.

**Fig. S2.**
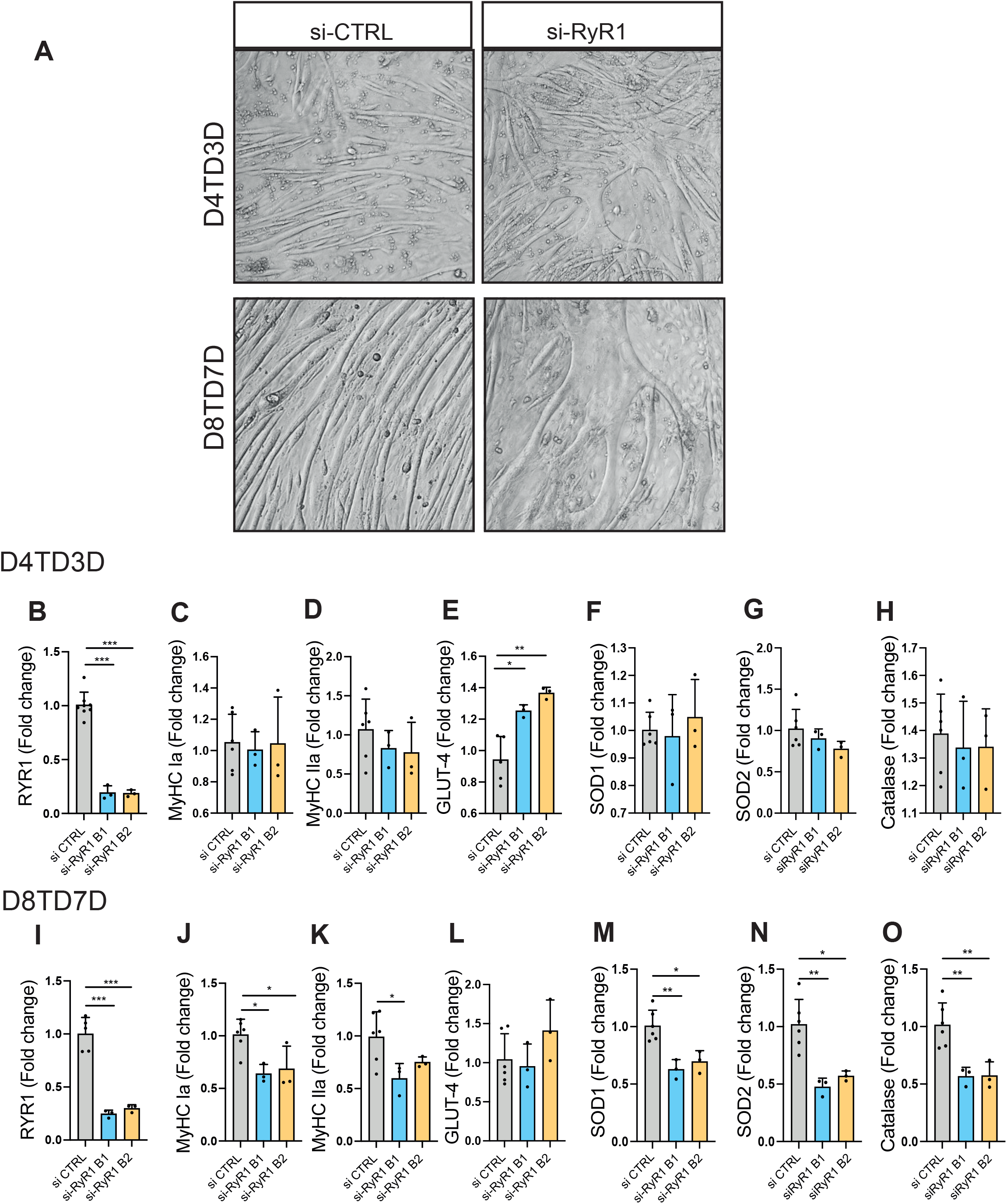
Characterization of si-RyR1 myotubes. (**A**) Brightfield images of si-CTRL and si-RyR1 myotubes at day 4 of transfection and day 3 of differentiation (D4TD3D) and day 7 of transfection and 6 of differentiation (D7TD6D) showing multi-sizes and disorganized myotubes in si-RyR1 cells as compared to controls. (**B-H**) Transcripts of RYR1, myosin heavy chain Ia (MyHC Ia), myosin heavy chain IIa (MyHC IIa), glucose transporter 4 (GLUT4, and antioxidant genes dismutase 1 (SOD1), dismutase 2 /SOD2) and catalase at D4TD3T, using two different batches of si-RNAs (B1 and B2). One-way ANOVA with Dunnett’s multiple comparisons test, n= 6, 3 and 3 independent biological experiments for si-CTRL, si-RyR1 B1 and si-RyR1 B2 respectively. (**I-O**) Transcripts of RYR1, myosin heavy chain Ia (MyHC Ia), myosin heavy chain IIa (MyHC IIa), glucose transporter 4 (GLUT4), and antioxidant genes dismutase 1 (SOD1), dismutase 2 /SOD2) and catalase at D7TD6T. One-way ANOVA with Dunnett’s multiple comparisons test, n= 6, 3 and 3 independent biological experiments for si-CTRL, si-RyR1 B1 and si-RyR1 B2 respectively *p ≤ 0.05, **p ≤ 0.01, ***p ≤ 0.001.

## Data availability

All data generated and analysed during this study are included in this published article (and its Supplementary information files). The proteomics data generated in this study have been deposited with PRIDE under accession code PXD039693.

## Author approval

All authors have seen and approved the manuscript.

## Acknowledgements

We thank Prof. Romano Regazzi and Prof. Dario Divani, directors of the Department of Biomedical Sciences for their support, infrastructure availability and discussions and Mr. Gilles Dubuis and Mr. Clément Lanfranchi for technical support. Special thanks to all the patients who provided their consent for the use of their muscle biopsy samples. We thank Prof. Isabelle Marty and Dr. Anne Petiot from the University of Grenoble for providing us with RyR1-Rec mice muscles and Prof. Andrew Marks from Columbia University for the RyR1 antibody. We also thank the proteomics, metabolomics and the cellular imaging facility platforms at the University of Lausanne. This work was supported by the SNF Grant (no. IZK0Z3_173941 to N.Z., PRIMA PR00P3_193166 to I.C.L.-M.) and the Subside “Tremplin/Relève Académique” of the UNIL to N.Z. J.V., B.K., N.P. and N.Z. were supported by institutional funds.

## Author contribution

Conceptualization: NZ

Methodology: M.W., PP.L, J.O., IC.LM., JA.L, N.P., N.Z.

Investigation: J.V., E.F., M.W., PP.L., AL.M, IC.LM., N.Z.

Visualization: M.W., PP.L., IC.LM., JA.L., B.K., N.P., N.Z.

Supervision: N.Z. Writing—original draft: N.Z.

Writing—review & editing: M.W., PP.L., J.O., IC.LM., JA.L., B.K., N.P., N.Z.

